# Calibration of transmission-dynamic infectious disease models: a scoping review and reporting framework

**DOI:** 10.1101/2025.03.09.25323613

**Authors:** Emmanuelle A. Dankwa, Léa Cavalli, Ruchita Balasubramanian, Melike Hazal Can, Hening Cui, Katherine M. Jia, Yunfei Li, Sylvia K. Ofori, Nicole A. Swartwood, Carrie Wade, Caroline O. Buckee, Jeffrey W. Imai-Eaton, Nicolas A. Menzies

## Abstract

**Objective/Background:** Transmission-dynamic models are commonly used to study infectious disease epidemiology. Calibration involves identifying model parameter values that align model outputs with observed data or other evidence. Inaccurate calibration and inconsistent reporting produce inference errors and limit reproducibility, compromising confidence in modeled results. No standardized framework exists for reporting on calibration of infectious disease models, and an understanding of current calibration approaches is lacking.

**Methods:** We developed a 15-item framework for reporting calibration practices and applied it in a scoping review to assess calibration approaches and evaluate reporting comprehensiveness in transmission-dynamic models of tuberculosis, HIV and malaria published between January 1, 2018, and January 16, 2024. We searched relevant databases and websites to identify eligible publications, including peer-reviewed studies where these models were calibrated to empirical data or published estimates.

**Results:** We identified 411 eligible studies encompassing 419 models, with 74% (n=309) being compartmental models and 20% (n=82) individual-based models (IBMs). The predominant analytical purpose was to evaluate interventions (71% of models, n=298). Parameters were calibrated mainly because they were unknown or ambiguous (40%, n=168), or because determining their value was relevant to the scientific question beyond being necessary to run the model (20%, n=85). The choice of calibration method was significantly associated with model structure (p-value<0.001) and stochasticity (p-value=0.006), with approximate Bayesian computation more frequently used with IBMs and Markov-Chain Monte Carlo with compartmental models. Regarding reporting comprehensiveness, all 15 framework items in the framework were reported in 4% (n=18) of models; 11-14 items in 66% (n=277), and 10 or fewer items in 28% (n= 124). Implementation code was the least reported, available in only 20% (n=82) of models.

**Conclusions:** Reporting on calibration is heterogeneous in recent infectious disease modeling literature. Our proposed framework for reporting of calibration approaches could support improved reproducibility and credibility of modeled analyses.

**Author Summary:** Calibration, the identification of parameter values so that model outcomes are consistent with observed data or other evidence, is often employed in the process of obtaining model results to inform health decision making. Despite its importance, there has not been a standardized framework for reporting how calibration is conducted in infectious disease modeling studies. This has led to inconsistent reporting practices and challenges in reproducing model results, potentially compromising confidence in their validity. We developed a calibration reporting framework, based on best practices found in the literature and informed by our expertise in conducting calibration. To assess calibration practices and their reporting, we applied our framework in a scoping review of 419 infectious disease transmission models of HIV, TB and malaria published between 2018 and 2024. Most models reviewed were compartmental (74%) or individual-based (20%), and the choice of calibration methods was associated with model structure and stochasticity. Calibration was conducted predominantly in the context of models aimed at evaluating the impact of disease control interventions, highlighting the role of calibration in decision making. Parameters were calibrated mainly because they were unknown or ambiguous, or because reporting their value was relevant to the scientific question beyond just being necessary to run the model. The comprehensiveness of calibration reporting varied across models, with most models omitting 1 to 5 items in the framework. Accessible implementation code was the most underreported, with only 20% of models including it. Our proposed framework could serve as a tool to standardize calibration reporting, thereby enhancing the transparency and reproducibility of calibration processes in transmission-dynamic models.

## 1 Introduction

Infectious disease models are designed to reproduce key features of disease epidemiology, including the impact of health services or interventions on the transmission or morbidity processes. These models are often used to predict disease trends or evaluate alternative interventions for disease control. Models are characterized by parameters: fixed values or variables that determine model behavior. Model calibration (or ‘model fitting’) includes a diverse range of methods for selecting values for model parameters such that the model yields estimates consistent with existing evidence. Specifically, a calibrated model is one in which the value of *at least one* parameter is chosen to achieve consistency of model outputs with empirical data, published estimates, or other evidence. In practice, this is commonly achieved by applying formal numerical optimization or statistical approaches that systematically vary parameters and assesses them according to a quantitative goodness-of-fit measure.

Calibration of infectious disease transmission models may be undertaken to infer the value of a parameter of epidemiological importance. These parameter estimates are interpreted as defining characteristics of the modeled diseases’ epidemiology, for example, the duration of the latent or incubation periods. Calibration can also be employed to enable the prediction of disease trends under a range of interventions. Such predictions are used as evidence to support policies on disease control; which attained widespread public prominence for the control of the COVID-19 pandemic (Brooks-Pollock et al., 2021; Centers for Disease Control and Prevention, 2020).

Inaccuracies in model calibration may result in inference errors, compromising the validity of modeled results that inform public health policies. Calibration is one among several choices that influence the validity of modeled evidence, such as model structure (Brisson and Edmunds, 2006), methodological assumptions (Brisson and Edmunds, 2006) and data type, all of which impact parameter identifiability (Dankwa et al., 2022; Eisenberg et al., 2013; Kao and Eisenberg, 2018; Tuncer and Le, 2018). The clarity, and therefore credibility, of model results may also be adversely influenced by an inadequate description of the calibration procedure employed in a study. This lack of thorough description and non-reporting of implementation code hampers reproducibility (Pokutnaya et al., 2023a), and potentially compromises trust in the validity of those studies for informing public health action.

Although calibration approaches are widely employed in infectious disease modeling, few studies have detailed their application, with most literature consisting of tutorials or best practices guidelines (Briggs et al., 2012; Chowell, 2017; Hazelbag et al., 2020; Jackson et al., 2015; Menzies et al., 2017; Vanni et al., 2011). In addition, efforts to develop a standardized framework for calibration have been limited. The Infectious Disease Modeling Reproducibility Checklist (IDMRC) (Pokutnaya et al., 2023a) provides a general framework for reporting modeling study components to ensure reproducibility but overlooks details specific to calibration, such as the uncertainty of calibration outputs (Ryckman et al., 2020) or the number of parameter sets calibrated. Hazelbag et al. (2020) proposed an extension to Stout et al.’s (2009) checklist for calibration reporting based on a review of individual-based transmission models (IBMs), but it has limited applicability to other model structures, like compartmental models. A standardized framework for calibration in infectious disease modeling could enhance objective and consistent reporting, ensuring that reports include all relevant details necessary for reproducibility. Moreover, no comprehensive overview exists of calibration approaches across all model types, examining how study context influences the choice of approach and evaluating reporting comprehensiveness. Understanding current calibration practices and their reporting could improve method selection and reporting and help identify areas for innovation and further research.

To address these gaps, we developed the *Purpose-Input-Process-Output (PIPO)* framework for reporting infectious disease model calibration. We applied this framework in a scoping review of the literature on transmission-dynamic models for tuberculosis (TB), HIV, and malaria published between January 1, 2018, and January 16, 2024. The review systematically mapped the conduct and reporting comprehensiveness of calibration in this field. It focused on evaluating the purpose, inputs, process, and outputs of calibration in recent literature, and examined how the choice of calibration approach varies by model type.

## 2 Methods

### 2.1 Purpose-Input-Process-Output (PIPO) framework

We developed the *Purpose-Input-Process-Output (PIPO) framework* as a proposed framework and checklist for reporting on infectious disease model calibration. This framework was developed based on the authors’ expertise in conducting calibration for transmission-dynamic models, and published guidance on calibration best practices (Briggs et al., 2012; Chowell, 2017; Hazelbag et al., 2020; Jackson et al., 2015; Menzies et al., 2017; Vanni et al., 2011).

PIPO is a 15-item reporting framework for describing calibration in infectious disease modeling studies to ensure reproducibility of calibration by facilitating a clear communication of calibration aims, methods and results. The framework has four broad components: 1) *Purpose*, which deals with the goal of calibration, 2) *Inputs*, which deal with the inputs into the calibration algorithm, 3) *Process*, which deals with how calibration is conducted, and 4) *Outputs*, which deals with the characteristics of the calibration outputs.

A brief overview of each component of the framework is as follows.

*Purpose*: This component collects information on the scientific problem being addressed by the study. For example, is the goal to understand disease mechanisms or to predict disease trends? Understanding the goal helps establish the context for the calibration.

*Inputs*: This component allows the reporting of key characteristics of the main inputs for calibration: 1) the parameters to be calibrated, and 2) the calibration targets. For parameters, PIPO includes fields to report on whether prior knowledge on any parameters is incorporated into the calibration process, whether all or just a subset of parameters was calibrated, and to justify why these parameters selected for calibration. For calibration targets, PIPO allows reporting on the number of calibration targets, the type of data used for defining targets (e.g., incidence, spatial data, etc.) and whether the targets are empirical data (as raw counts or numbers and their corresponding statistical summaries, such as means, medians, rates, or proportions), or modeled estimates. The absence of specific information on parameters and calibration targets when reproducing a calibration result has many implications. For example, parameter estimates obtained through calibration may differ depending on which parameters are fixed (held at a constant value in the model) and which are calibrated. Also, the type of data used as a calibration target could affect calibrated parameter estimates. For instance, the identifiability of parameters may vary depending on whether incidence, prevalence or another type of data is used as calibration target (Dankwa et al., 2022). Therefore, without clear reporting on the characteristics of calibration targets and parameters, the reproducibility of calibration and model results is hampered.

*Process*: This component allows the reporting of all details related to the calibration method used. Reporting covers the name of the calibration method and a brief description of how it works, including its goodness-of-fit (GOF) measure, which assesses the level of agreement between modeled outcomes and calibration targets. Further, the component allows reporting the following details of the calibration method’s implementation: 1) a well-documented, accessible repository for calibration code (if used), 2) programming language, and 3) versions for any programming languages, packages, or data repository used. In our review of models, we examined the availability of any model implementation code, acknowledging that calibration code is not typically reported separately.

*Output*: Characteristics of calibrated parameter estimates and the corresponding model outcomes, jointly termed “calibration outputs” are reported in this component. Reporting items include specification of whether the parameter values produced by calibration represented a “point estimate (i.e., a single parameter or parameter set), a “sample estimate” (i.e., multiple parameter values or sets) or a “distribution estimate” (i.e., a closed-form distribution function that could be used to generate new parameter values), as can be obtained with Laplace approximation (e.g., (Maheu-Giroux et al., 2019). Details on the size of calibration outputs and uncertainty in the output are also reported here, as they are relevant for reproducibility. For calibration processes that produce a sample of parameter values or sets, an insufficient sample size can introduce additional uncertainty into modelled outcomes. For this reason, the sample size of calibrated parameters used for inferences must be clearly reported.

The full framework is available in the supplementary material and has been formatted such that it is readily available for use as a checklist for calibration reporting.

### 2.2 Scoping Review

#### 2.2.1 Review conduct and reporting

We reported the review following the Preferred Reporting Items for Systematic Review and Meta-analysis Extension for Scoping Reviews (PRISMA-ScR) checklist (Tricco et al., 2018) (Table S2). In developing the protocol and conducting the review, we followed the guidelines proposed by Peters et al. (2020). The review protocol was pre-registered with the Open Science Framework (https://doi.org/10.17605/OSF.IO/3VTJW).

#### 2.2.2 Eligibility criteria and information sources

We used the Studies, Data, Methods and Outcomes (SDMO) Framework (Munn et al., 2018b) to define the inclusion and exclusion criteria based on the research question: In transmission-dynamic models of TB, HIV and malaria, how is calibration conducted? Inclusion and exclusion criteria are detailed in Table S1, and summarized as follows: Published and peer-reviewed studies in which a transmission-dynamic model for HIV, TB or malaria was calibrated to empirical data or published estimates were included. Following Pitman et al. (2012), we defined a dynamic infectious disease transmission model as a mathematical model for describing infection or disease spread, where the risk of infection is not constant and depends on the number of infectious individuals in the population at any given time, thus allowing for the modeling of nonlinear feedback effects. We focused on dynamic models, acknowledging that the norms of calibration of dynamic models may differ from those of other models, such as static models, which make the assumption that the risk of infection is constant over time (Porgo et al., 2019). We defined calibration as the use of a method to select values or distributions for model parameters so that model outputs were consistent with observed data or estimates, referred to as *calibration targets*. Unpublished studies, studies from proceedings, opinion pieces or animal studies were excluded. We also excluded studies that were not published in the English language due to limitations with translating and interpreting foreign language studies. To focus our review on current practices, we restricted our search to studies published within approximately 5 years of the search date, covering the period 1 January 2018 to 16 January 2024 (the search date). Table S3 describes the full search strategy, which was constructed with librarian support [CW]. Briefly, we searched PubMed, Embase, lobal Health, Web of Science Core Collection, and Global Index Medicus databases to identify eligible studies that contain in their title or abstract keywords related to “transmission-dynamic models”, and “HIV”, “tuberculosis” or “malaria”. We also searched relevant websites of major disease consortia for HIV (HIV Modeling Consortium: http://hivmodeling.org/) and TB (TB Modelling and Analysis Consortium: https://tb-mac.org/). Although we identified a consortium for malaria vaccine models, we did not add this to our search because we sought for collections that were general and not topic specific.

#### 2.2.3 Title and abstract screening

Before title and abstract screening, the screening review team (EAD, LC, RB, MHC, KMJ and SKO) pilot tested the inclusion/exclusion criteria as follows: A random sample of 14 titles/abstracts were selected and each team member independently screened these using the eligibility criteria. An agreement of 85% was obtained. The team then discussed the discrepancies and modified the criteria as needed. For title/abstract screening, we applied all inclusion and exclusion criteria except the criterion that studies needed to involve calibration. This is because calibration may not always be mentioned in the abstract even if it was performed in a study. Each title and abstract were reviewed independently by two reviewers, with conflicts adjudicated by a third reviewer.

#### 2.2.4 Full text screening

Studies not screened out based on their titles and abstracts underwent full text screening. Most full texts were either freely available online or accessible via institutional library access. We excluded studies for which full texts were not accessible. Each full text was read independently by two reviewers, and the full inclusion and exclusion criteria were applied. Conflicts were resolved by a third reviewer.

#### 2.2.5 Data extraction

We extracted studies using the PIPO framework. The data extraction form for this review was expanded to additionally include general study characteristics such as the year of publication, disease of focus and model structure. The form had 22 reporting items in total: 15 items corresponding to the PIPO framework and 7 items on general study characteristics. The form was developed with input from all authors and tested by a team of four reviewers (EAD, LC, RB, KMJ). After incorporating feedback from the first round of testing, two reviewers (EAD and LC) conducted a second instance of testing. The final form is available as supplementary material. For each study, data were independently extracted by two reviewers in the data extraction team (EAD, LC, RB, MHC, YL, NS, HC). Conflicts were resolved by a third reviewer in the team.

#### 2.2.6 Evaluating the conduct of calibration in the recent literature

First, we summarized the extracted data as counts and percentages for every extracted item. Additionally, we performed statistical tests to assess the relationship between choice of calibration method, model structure and model stochasticity. To ensure large sample sizes and sufficient statistical power, statistical tests were limited to calibration methods that were used in at least 30 models and to model structure or stochasticity categories that had at least one report for each of these calibration methods. For the test involving model stochasticity, we employed a chi-square test. For the test involving model structure, we used the Fisher’s exact test as expected values were small for some cells hence the Chi-square approximation, which assumes large samples, could not be used. Logistic regression was used to determine the relationship between calibration methods and model structure and stochasticity. The figures of test results were produced using the “ggstatsplot” package (v 0.13.0) (Patil, 2021) in R (v 4.3.1).

#### 2.2.7 Evaluating the comprehensiveness of calibration reporting

To investigate the extent to which calibration reporting was comprehensive, we defined a 15-item comprehensiveness scale, corresponding to items in the PIPO reporting framework. For each study, we assigned a score of 1 if an item was reported or a score of 0 if it was not reported. Therefore, the maximum attainable score was 15, indicating a study with high reporting comprehensiveness, and the minimum attainable score was 0, indicating a study with low reporting comprehensiveness. We categorized studies by score as follows. A study with a score of 0-5 was categorized as having “low” reporting comprehensiveness; a study with a score of 6-10 was categorized as having “fair” reporting comprehensiveness, and a study with a score of 11-15 was categorized as having “high” reporting comprehensiveness. We interpreted reporting comprehensiveness as indicative of transparency around calibration methods and potential for reproducibility.

#### 2.2.8 Analysis tools

Screening and data extraction were performed in Covidence (Veritas Health Innovation, 2024). Analyses of extracted data were performed in R version 4.3.1. (R Core Team, 2024).

#### 2.2.9 Reproducibility

The full, commented analysis code and relevant data dependencies are available at the repository: https://github.com/Leacavalli/Calibration-Review.

#### 2.2.10 Results validation

One reviewer (LC) conducted data cleaning on the raw extracted data to address inconsistencies due to varying reporting styles across studies and reviewers. This included standardizing free-text entries that were reported differently. The data cleaning code is available on GitHub. In addition, when inconsistencies in the extracted data were suspected during analysis, two reviewers (LC and EAD) checked the relevant sections of the extracted data and where necessary, also checked with the original data extractor to ensure accuracy.

## 3 Results

### 3.1 Study selection

The literature search yielded 6518 studies. After duplicates were removed, 3138 studies remained to be screened by title and abstract. Of these, 765 progressed to the full text review stage. We excluded 354 of these studies based on the exclusion criteria or because they did not have accessible full texts (n=4). We included 411 studies in the review (Figure 1).

**Figure 1:**
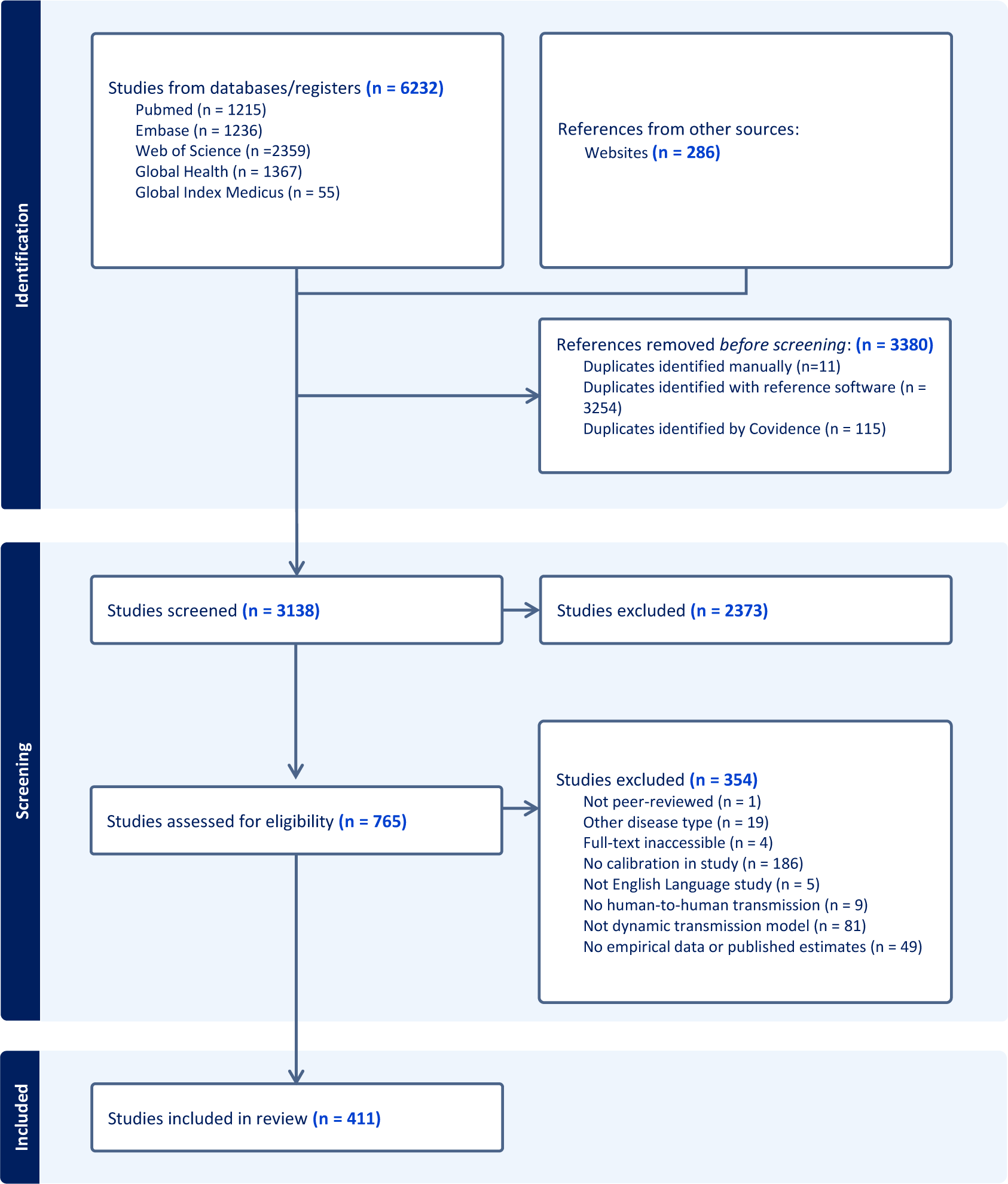
PRISMA flow chart of study selection process.

### 3.2 General model characteristics

The 411 included studies yielded 419 unique model applications and their calibrations, as five studies calibrated multiple models (Table S4). The full extracted data for each calibrated model are in Sheet S1. Summary statistics (i.e., counts and percentages, where applicable) for each extracted item are in Sheet S2.

Table 1 summarizes the general characteristics of all 419 calibrated models. Of the models studied, 48% (n=203) focused exclusively on HIV, 33% (n=138) on TB, 16% (n=67) on malaria, and 3% (n=11) on combined HIV/TB. In structure, models were mostly compartmental (74%, n=309) or individual-based (20%, n=82). One model (Unwin et al., 2021) used a Hawkes process. Model structure was unreported in 6% (n=23) of the models. Deterministic models accounted for 36% (n=149) of reviewed models, while stochastic models accounted for 22% (n=91). Whether or not a model included stochasticity was unreported in the remaining 43% (n=179).

**Table 1:**
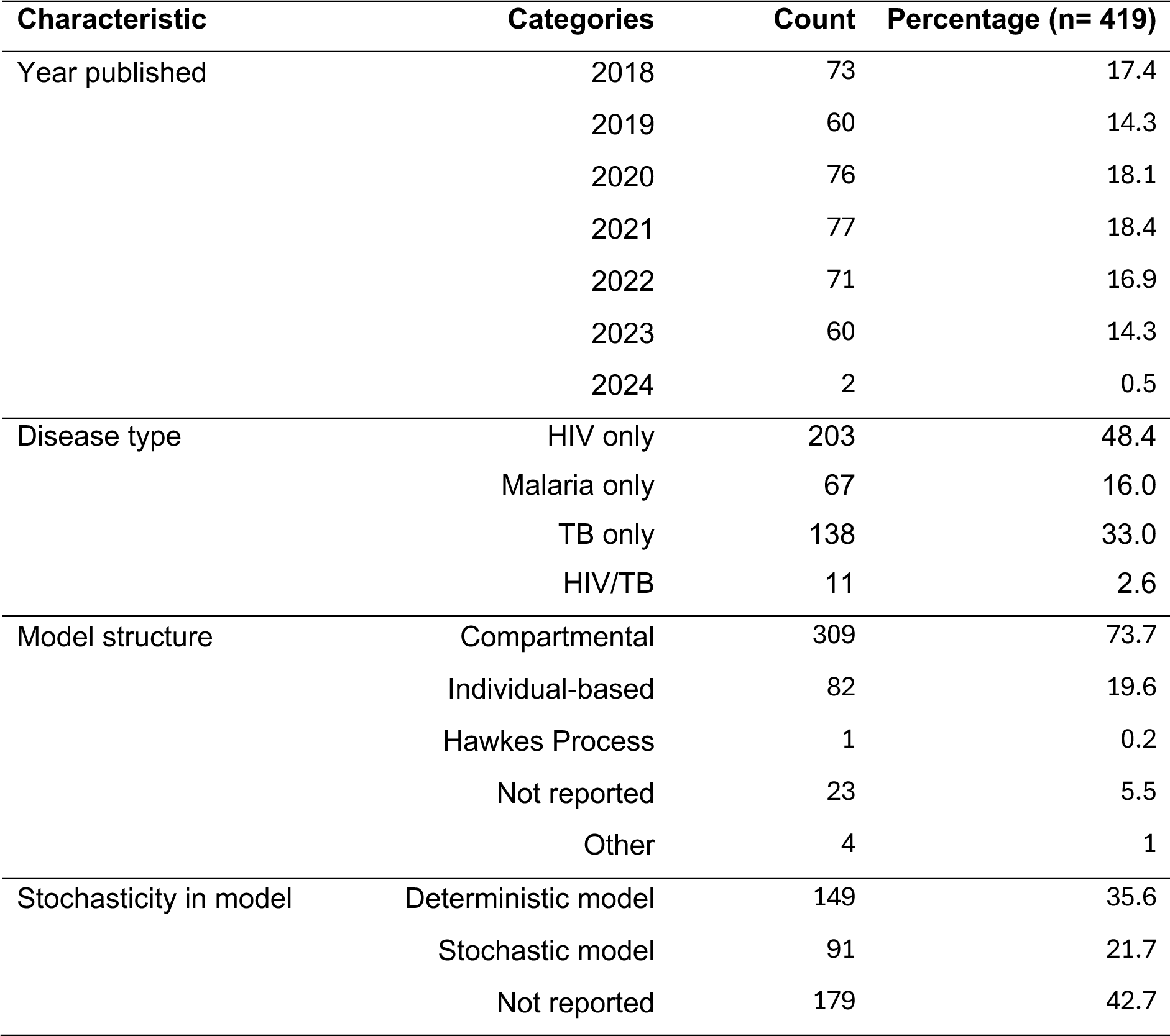
General characteristics of calibrated models (n=419).

### 3.3 Conduct of calibration in the literature

#### 3.3.1 Purpose

Predominantly, calibration was conducted in the context of evaluating or comparing interventions (71% of models, n =298). Other purposes were to understand disease mechanisms (38%, n=161) or predict disease trends (24%, n=102). The least frequent purpose of calibration was to assess the impact of model assumptions (9%, n=37). Some models (40%, n=169) were calibrated for multiple reasons (therefore, percentages add to >100%).

#### 3.3.2 Inputs

For most models (63%, n=263), prior knowledge about parameters to be calibrated was incorporated in the calibration process. In many cases, this prior knowledge was obtained from the literature, evidenced by citations of other papers for parameter sources. In most models (92%, n=384), calibration was limited to a subset of model parameters. Only 12 models (3%), where reported, had all parameters calibrated. Parameters were selected for calibration because they were unknown or ambiguous (40%, n=168), and/or because determining and reporting their accurate value was relevant to the question of interest beyond just being necessary to run the model (20%, n=85). Most model reports (45%, n=190) did not include a justification for the choice of parameters to be calibrated.

The main types of data used for defining calibration targets were disease prevalence (48% of models, n=201), notifications or diagnoses (46%, n=193), incidence (42%, n=176), treatment-related data (39%, n=165) and demographic data (28%, n=118) (Figure S1). The least used data types for calibration were spatial data (n=3), cost data (n=1) and effect sizes from trials (n=1). Calibration targets were mostly empirical data or their corresponding statistical summaries (86%, n=359), while the use of modelled estimates was less common (34%, n=142). Most models were calibrated to multiple targets (72%, n=300) rather than a single calibration target (26%, n=110). In 2.1% (n=9) of models, the number of calibration targets was not reported.

#### 3.3.3 Process: Calibration methods

The four most frequently used calibration methods were least squares estimation (20%, n=83), Markov Chain Monte Carlo (MCMC) methods (16%, n=67), Approximate Bayesian Computation (ABC) (10%, n=40) and maximum likelihood estimation (8%, n=32). Each of these methods were employed in at least 30 models. In 20 models (5%), parameters were calibrated manually without relying on a numerical optimisation algorithm, i.e., by hand-tuning.

We classified calibration methods into two broad families according to the aim of the calibration procedure, inferred from the nature of the calibration results as extracted from a study (see item D.1. in the PIPO framework). If the nature of the calibration result was a “Sample estimate” or “Distribution estimate”, we concluded that the aim of the procedure was to approximate a distribution. If the nature of the calibration result was a “Point estimate”, we concluded that the aim of the procedure was to identify a single optimal parameter set. For studies for which the nature of the calibration result was not clear or judged as “Other”, we revisited the study for more details to determine the most appropriate aim of the calibration procedure. We reviewed 18 models aiming to identify an optimal parameter set and 7 aiming to approximate a distribution (Table 2). A brief description for each calibration method we reviewed is provided in Supplementary Table S5.

**Table 2:** Calibration methods classified by aim of calibration procedure. The number and percentage of models applying these methods have also been indicated. Brief descriptions of methods are in Supplementary Table S5.

| Calibration method | Number of models | Percentage of models (n=419) |
| --- | --- | --- |
| **Aim: Approximate a distribution** |  |  |
| Markov Chain Monte Carlo (MCMC) | 67 | 16.0 |
| Approximate Bayesian Computation (ABC) | 40 | 9.5 |
| Incremental Mixture Importance Sampling (IMIS) | 15 | 3.6 |
| Sequential Monte Carlo (SMC) | 12 | 2.9 |
| Sampling importance resampling (SIR) | 9 | 2.1 |
| History matching with emulation (HME) | 7 | 1.7 |
| Laplace Approximation | 1 | 0.2 |
| **Aim: Identify an optimal parameter set** |  |  |
| Least squares estimation | 83 | 19.8 |
| Maximum likelihood estimation | 32 | 7.6 |
| Manual (hand-tuning) | 20 | 4.8 |
| Nelder-Mead algorithm | 6 | 1.4 |
| Grey Wolf Optimizer (GWO) | 3 | 0.7 |
| Genetic algorithm | 2 | 0.5 |
| Parallel Simultaneous Perturbation Optimisation (PSPO) | 2 | 0.5 |
| Broyden-Fletcher-Goldfarb-Shanno algorithm | 2 | 0.5 |
| Coordinate descent algorithm | 1 | 0.2 |
| Deviance-based loss | 1 | 0.2 |
| Levenberg-Marquardt optimization algorithm | 1 | 0.2 |
| Local sequential quadratic programming (DESQP) optimization algorithm | 1 | 0.2 |
| Maximum-a-Posteriori estimation | 1 | 0.2 |
| Mean square error with threshold | 1 | 0.2 |
| Method of moments | 1 | 0.2 |
| Minimum Step Deviation | 1 | 0.2 |
| Minimum contrast estimation | 1 | 0.2 |
| Root Mean Squared (RMS) deviation | 1 | 0.2 |

The choice of calibration method was significantly associated with type of model structure (p-value < 0.001, Figure S2) and model stochasticity (p-value = 0.006, Figure S3). The ABC method was used significantly more frequently with IBMs than with compartmental models (odds ratio (OR)= 7.7, 95% CI: 3.4-17.8, p<0.001), with 50% of IBMs (n=17/34) and 12% of compartmental models (n=20/175) being calibrated using ABC (Figure S2). Conversely, MCMC methods were used significantly more frequently with compartmental models than with IBMs (OR=3.3, 95% CI: 1.3-12.6, p=0.021), with 12% of IBMs (n=4/34) and 33% of compartmental models (n=57/175) being calibrated using MCMC (Figure S3). The odds of using least square and maximum likelihood estimation did not significantly differ by model structure (respectively: OR=0.5, 95% CI: 0.2-1.1, p= 0.10, and OR= 0.8, 95% CI: 0.2-2.2, p=0.70). The association patterns of calibration methods with model stochasticity were similar, with the ABC method being significantly more common with stochastic models (OR=4.2, 95% CI: 1.8-10.4, p=0.001), while the MCMC method was more frequently used with deterministic models, although this relationship was not statistically significant (OR=2.4, 95% CI: 1.0-5.8, p=0.051). The least square and maximum likelihood estimation methods showed no significant association with stochasticity (LSE: OR=0.6, 95% CI: 0.3-1.4, p=0.24; MLE: OR=1.1, 95% CI: 0.4-2.9, p=0.91). This aligns with the tight link between model structure and stochasticity, since 88% of compartmental models with reported stochasticity were deterministic (n=80/91), while 97% of IBMs (n=30/31) were stochastic.

#### 3.3.4 Process: Goodness-of-fit (GOF) measures

The GOF measures employed as part of the calibration process were based on an ad-hoc distance function (27%, n=115), data likelihood (22%, n=93) or another measure (5%, n=22), such as the deviance information criterion (e.g., Shaweno et al.,(2018)), and the Akaike information criterion (e.g., Kim et al.,(2020), Mettle et al. (2020), and Mussina et al.,(2023)). For 45% of models (n=189), the GOF measure was either not reported at all or not reported sufficiently clearly to allow extraction of relevant information.

#### 3.3.5 Process: Software

Calibration implementation code was reported in an open-access repository for only 20% of models (n=82). For 9 models (2%), the calibration code was inaccessible although a link to a repository was reported. For most models (78%, n=328), calibration code was not reported at all. Regarding programming languages used for calibration, the following were the most used: R (26%, n=110), MATLAB (20%, n=84), C++ (8%, n=33) and Python (6%, n=23), in order of frequency of use. Notably, for 181 models (43%, n=181), the programming language used was not reported. Multiple programming languages were used in 33 models (8%).

#### 3.3.6 Output

Where reported, calibration outputs were typically point estimates (41.5%, n=174) or a “sample estimate” (i.e., multiple parameter values or sets, 45%, n=189). Only one model (Maheu-Giroux et al., 2019) presented results as a “distribution estimate”. In this model, calibration was achieved through a Laplace approximation process, which provided a closed-form approximation of the posterior distributions for both parameter values and model outputs. Reporting of calibration outputs was exclusively numerical (10%, n=40), exclusively graphical (13%, n=53) or, in most cases (74%, n=311), a combination of numerical and graphical approaches. Calibration outputs were not reported at all in 15 models (4%). Regarding uncertainty in parameter estimates, we observed both numerical (i.e., as confidence or credible intervals) and graphical (i.e., shaded areas around curves on graphs) reporting in 41% (n=170) of models, exclusive numerical reporting in 12% (n=52) and exclusive graphical reporting in 15% (n=64). Many model reports (32%, n=133) did not report on the uncertainty in their parameter estimates.

Among calibration processes that generated a multiple sets of parameter values (i.e. a “sample estimate”), there was substantial variation in the size of calibration outputs (Figure S4). The largest calibration outputs, with over 2000 parameter sets, were only observed with MCMC, ABC, and Incremental Mixture Importance Sampling.

#### 3.3.7 Comprehensiveness of calibration reporting

The least reported item in the PIPO framework was the implementation code, which was reported and accessible in only 20% of models (n=82). Other less reported items—reported in fewer than 70% of models—were the justification for the choice of parameters to calibrate (55%, n=229), whether calibration was done in a single step or sequentially (56%, n=234), the GOF measure used within the calibration process (57%, n=238), whether external beliefs or evidence was used for calibration (66%, n=276) and the uncertainty in parameter estimates (68%, n=286).

Items with high reporting frequencies were the type (99%, n=416) and resolution (95%, n=397) of data used for defining calibration targets, the number of calibration targets (98%, n=410), calibration outputs (96%, n=404) and the choice of parameters to calibrate (95%, n=396).

The reporting comprehensiveness was excellent (all 15 items reported) in 4% of models (n=18), high (11-14 items reported) in 66% of models (n=277), fair (6-10 items reported) in 28% (n=118) and low (0-5 items reported) in 1% (n=6) (Figure 2). We did not observe differences in the distributions of reporting comprehensiveness by year of model publication (Figure S5). Individual reporting item scores for all 419 models are presented in Sheet S3.

**Figure 2:**
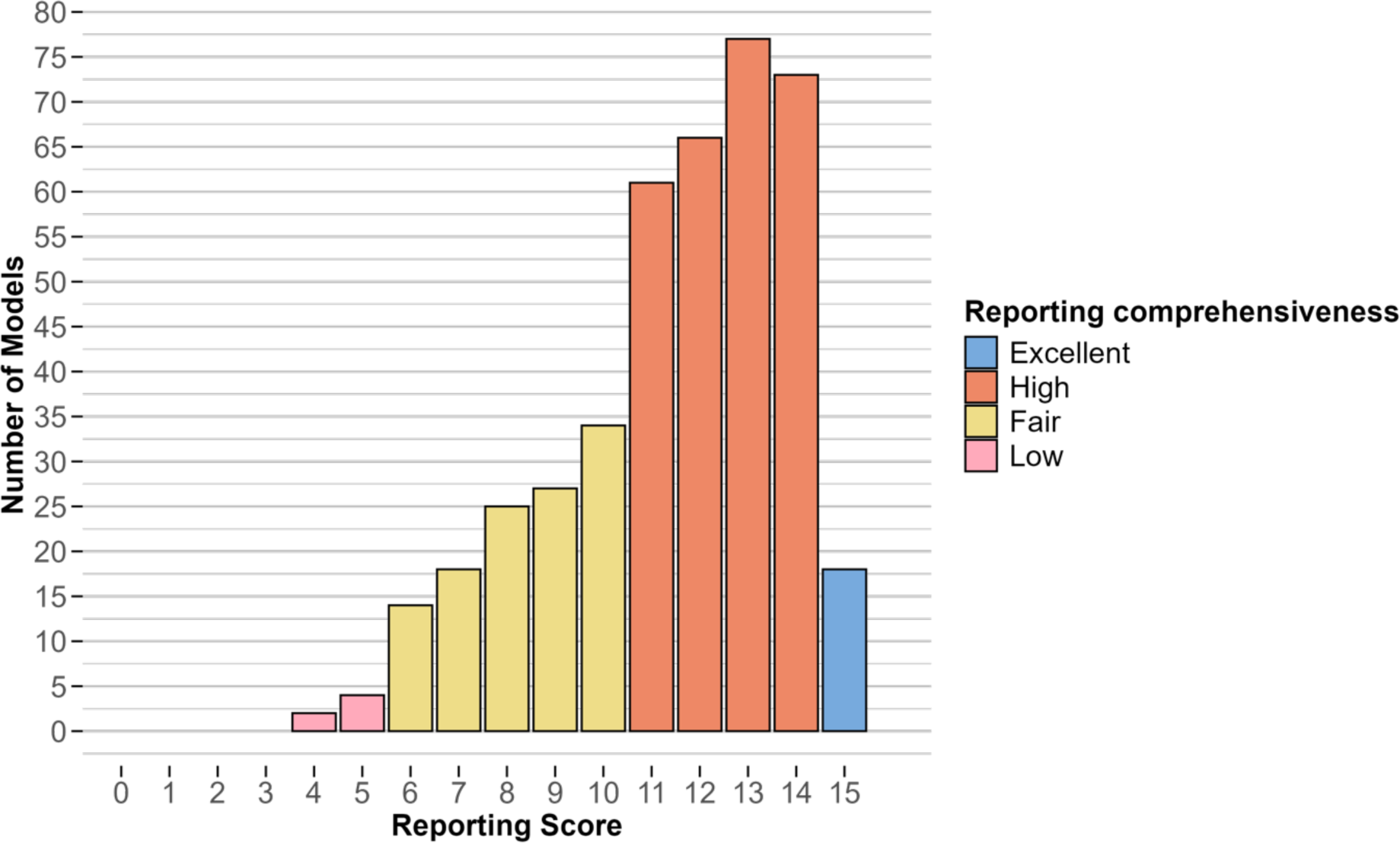
Distribution of scores for calibration reporting comprehensiveness for 419 models.

## 4 Discussion

We developed a 15-item framework for calibration reporting, encompassing criteria regarding the purpose, inputs, process, and outputs (PIPO) of calibration, based on best practices on calibration in the literature, and the authors’ expertise in conducting calibration. Our PIPO framework addresses the shortcomings of previous reporting frameworks (Hazelbag et al., 2020; Pokutnaya et al., 2023a) in several ways. Specifically, PIPO is applicable to all model structures, whereas the framework in Hazelbag et al. (2020) was primarily designed for IBMs. Additionally, PIPO includes a field for code reporting, which is crucial for ensuring reproducibility. We recommend using the PIPO framework as a complement to the IDRMC, as it specifically expands on elements specific to calibration reporting not captured in the IDMRC, such as the uncertainty and size of calibration outputs, which are essential for model reproducibility.

We applied the PIPO framework to examine current calibration conduct and reporting practices through a scoping review of 419 models from 411 studies on HIV, TB, and malaria transmission-dynamic modelling, published between 2018 and 2024. Our review showed that in 30% of models (n=124), fewer than 11 items on the PIPO framework were reported, with the lowest reporting observed for calibration implementation code, reported in only 20% of models (n=82). This is lower than was observed in a recent review of early COVID-19 modeling studies (Pokutnaya et al., 2023b). Efforts to promote efficient code reporting, such as the development of tools and best practice guidelines (Piccolo and Frampton, 2016) and including reproducible software development (Gallagher et al., 2024) in the training of modelers should be encouraged.

We observed that a large fraction of model reports (45%, n=190) did not justify the selection of specific parameters for calibration over others. Explicitly considering whether a parameter needs to be calibrated prompts important questions including whether calibration targets are appropriate for informing the parameter’s value/distribution, whether alternative data on the parameter exists, and whether calibration will be relevant for addressing the modelling problem. When considered earlier in the study process, such questions could promote efficiency by reducing time spent exploring calibration avenues that may not be feasible given the study design and available data. In line with the findings of (Hazelbag et al., 2020), many models in our review (45%, n=189) did not report or were unclear about the GOF measure used. Given that the GOF measure evaluates the level of agreement between modeled outcomes and calibration targets, it is essential to define and report the GOF measure clearly and quantitatively, rather than relying on subjective or non-quantitative measures, thereby ensuring reproducibility and credibility of studies.

Most models (86%, n=359) relied on empirical data or their summaries—examples including prevalence and notifications—for defining calibration targets (Figure S1). Because of this reliance on empirical data, efforts to improve its quality and completeness should be emphasized. While not captured by the PIPO framework, we encourage authors to evaluate the quality of empirical data prior to its use, as it could directly influence the quality of parameter estimates. Efforts to improve data collection systems and the quality of collected data will not only aid in infectious disease surveillance, but also in the development of more accurate calibrated models. Furthermore, while not evaluated in our review, potential errors or missingness in the data also needs to be accounted for or corrected prior to data use for calibration to minimize biases in estimates. When modeled estimates rather than empirical data are used to define calibration targets, it is important to consider how the underlying uncertainty in these targets could be reflected in the uncertainty in parameter estimates. For some of the models we reviewed (68%, n=286), uncertainty estimates for their calibration outputs were reported but we did not study the sources of this uncertainty. A potential area of future work is to explore how uncertainty in calibration targets, as well as other types of uncertainty (model structure uncertainty, stochastic uncertainty) are captured in uncertainty estimates for parameters.

The choice of calibration method varied with the type of model structure, based on statistical tests of independence. The ABC method, for example, was used much more frequently with IBMs than with compartmental models (Figure S2). These differences can be explained in part by the compatibility between model structure and calibration method. Compared to compartmental models, IBMs are more complex in structure and therefore tend to have more complex likelihood functions that are difficult to specify. Given that ABC allows for likelihood-free inference (Beaumont et al., 2002), it is suitable for use with IBMs for which a likelihood function may be unavailable. The significant relationship between model stochasticity and calibration method could similarly be explained, at least in part, by model complexity. For instance, complex IBMs, usually associated with likelihood-free calibration methods like ABC (Figure S2), are also typically stochastic (Willem et al., 2017). This co-occurrence may explain why, among stochastic models, we observed a greater preference for methods such as ABC and a lower preference for likelihood-dependent methods such as MCMC (Figure S3).

This review has several limitations. First, the calibration practices examined here may have limited generalizability beyond models with structures like those used for HIV, malaria and TB and identified in the review. These diseases were selected because they are among the most extensively studied epidemics, contributing significantly to the global burden of disease. They also encompass a diverse range of transmission routes and natural history patterns, and thus a variety of model types. Nevertheless, their coverage is not exhaustive. Alternative transmission routes, such as the fecal-oral route exhibited by cholera and other gastrointestinal illness, are missing. Given that transmission routes and natural history influence model structure, we suspect that some model types are not represented. Second, there is a potential language bias, as the review was limited to studies published in the English language, such that we may have missed model types that only tend to be published in other languages. Third, we did not track repeated uses of the same core model, which could have potentially biased our results to reflect characteristics of models that are more generalizable or adaptable.

In conclusion, we have evaluated the conduct and reporting of calibration in 419 recently published infectious disease transmission models. In this review, the conduct and reporting of calibration was found to be highly heterogeneous, indicating a potential role for a standardized reporting framework to enhance reproducibility of results and consequently, the credibility of model results used for health decision making. Based on information from the reviewed models, best practices in literature and authors’ expertise in calibration, we developed the PIPO calibration reporting framework that captures four important components of calibration: its purpose, inputs, process and outputs. Use of this framework as a reporting tool could enhance standards in calibration reporting and improve the credibility and reproducibility of infectious disease model results. Moving forward, it is essential to continually monitor potential shift in the types of methods and adapt calibration methods and reporting standards accordingly.

## Supporting information

Sheet S1

Sheet S2

Sheet S3

Data extraction form

PIPO reporting framework

## Data Availability statement

All data relevant to this study are included or linked in the article.

## Acknowledgements

None

## Author contributions

EAD conceived the study. EAD and NAM designed the study and developed the review protocol. JWI-E and CB refined the protocol. CW, EAD and NAM developed the search strategy. CW and EAD ran the search. EAD, LC, RB and KMJ validated the data extraction form. EAD, LC, RB, KMJ, MHC, SKO, NAS, YL and HC identified eligible studies and extracted data. LC conducted data cleaning. EAD and LC performed the data analysis and validation. EAD wrote the first manuscript draft. All authors reviewed and edited the manuscript.

## References

Beaumont, M.A., Zhang, W., Balding, D.J., 2002. Approximate Bayesian computation in population genetics. Genetics 162, 2025–2035.

Briggs, A.H., Weinstein, M.C., Fenwick, E.A.L., Karnon, J., Sculpher, M.J., Paltiel, A.D., 2012. Model Parameter Estimation and Uncertainty: A Report of the ISPOR-SMDM Modeling Good Research Practices Task Force-6. Value in Health 15, 835–842. 10.1016/j.jval.2012.04.014

Brisson, M., Edmunds, W.J., 2006. Impact of Model, Methodological, and Parameter Uncertainty in the Economic Analysis of Vaccination Programs. Med Decis Making 26, 434–446. 10.1177/0272989X06290485

Brooks-Pollock, E., Danon, L., Jombart, T., Pellis, L., 2021. Modelling that shaped the early COVID-19 pandemic response in the UK. Philos Trans R Soc Lond B Biol Sci 376, 20210001. 10.1098/rstb.2021.0001

Centers for Disease Control and Prevention, 2020. COVID-19 Forecasting and Mathematical Modeling [WWW Document]. Centers for Disease Control and Prevention. URL https://www.cdc.gov/coronavirus/2019-ncov/science/forecasting/forecasting-math-modeling.html (accessed 12.15.23).

Chowell, G., 2017. Fitting dynamic models to epidemic outbreaks with quantified uncertainty: A primer for parameter uncertainty, identifiability, and forecasts. Infect Dis Model 2, 379–398. 10.1016/j.idm.2017.08.001

Dankwa, E.A., Brouwer, A.F., Donnelly, C.A., 2022. Structural identifiability of compartmental models for infectious disease transmission is influenced by data type. Epidemics 41, 100643. 10.1016/j.epidem.2022.100643

Eisenberg, M.C., Robertson, S.L., Tien, J.H., 2013. Identifiability and estimation of multiple transmission pathways in cholera and waterborne disease. Journal of theoretical biology 324, 84–102.

Gallagher, K., Creswell, R., Lambert, B., Robinson, M., Lei, C.L., Mirams, G.R., Gavaghan, D.J., 2024. Ten simple rules for training scientists to make better software. PLOS Computational Biology 20, e1012410. 10.1371/journal.pcbi.1012410

Hazelbag, C.M., Dushoff, J., Dominic, E.M., Mthombothi, Z.E., Delva, W., 2020. Calibration of individual-based models to epidemiological data: A systematic review. PLOS Computational Biology 16, e1007893. 10.1371/journal.pcbi.1007893

Jackson, C.H., Jit, M., Sharples, L.D., De Angelis, D., 2015. Calibration of complex models through Bayesian evidence synthesis: a demonstration and tutorial. Med Decis Making 35, 148–161. 10.1177/0272989X13493143

Kao, Y.-H., Eisenberg, M.C., 2018. Practical unidentifiability of a simple vector-borne disease model: Implications for parameter estimation and intervention assessment. Epidemics 25, 89–100.

Kim, S., Byun, J., Park, A., Jung, I., 2020. A mathematical model for assessing the effectiveness of controlling relapse in Plasmodium vivax malaria endemic in the Republic of Korea. PLOS ONE 15. 10.1371/journal.pone.0227919

Maheu-Giroux, M., Marsh, K., Doyle, C.M., Godin, A., Lanièce Delaunay, C., Johnson, L.F., Jahn, A., Abo, K., Mbofana, F., Boily, M.-C., Buckeridge, D.L., Hankins, C.A., Eaton, J.W., 2019. National HIV testing and diagnosis coverage in sub-Saharan Africa: a new modeling tool for estimating the ‘first 90’ from program and survey data. AIDS 33, S255. 10.1097/QAD.0000000000002386

Menzies, N.A., Soeteman, D.I., Pandya, A., Kim, J.J., 2017. Bayesian Methods for Calibrating Health Policy Models: A Tutorial. Pharmacoeconomics 35, 613–624. 10.1007/s40273-017-0494-4

Mettle, F.O., Osei Affi, P., Twumasi, C., 2020. Modelling the Transmission Dynamics of Tuberculosis in the Ashanti Region of Ghana. Interdisciplinary Perspectives on Infectious Diseases 2020, 4513854. 10.1155/2020/4513854

Munn, Z., Peters, M.D.J., Stern, C., Tufanaru, C., McArthur, A., Aromataris, E., 2018a. Systematic review or scoping review? Guidance for authors when choosing between a systematic or scoping review approach. BMC Medical Research Methodology 18, 143. 10.1186/s12874-018-0611-x

Munn, Z., Stern, C., Aromataris, E., Lockwood, C., Jordan, Z., 2018b. What kind of systematic review should I conduct? A proposed typology and guidance for systematic reviewers in the medical and health sciences. BMC Med Res Methodol 18, 1–9. 10.1186/s12874-017-0468-4

Mussina, K., Kadyrov, S., Kashkynbayev, A., Yerdessov, S., Zhakhina, G., Sakko, Y., Zollanvari, A., Gaipov, A., 2023. Prevalence of HIV in Kazakhstan 2010-2020 and Its Forecasting for the Next 10 Years. HIV 15, 387–397. 10.2147/HIV.S413876

Patil, I., 2021. Visualizations with statistical details: The “ggstatsplot” approach. Journal of Open Source Software 6, 3167. 10.21105/joss.03167

Peters, M., Godfrey, C., McInerney, P., Munn, Z., Tricco, A., Khalil, H., 2020. Chapter 11: Scoping reviews (2020 version), in: Aromataris, E., Munn, Z. (Eds.), JBI Manual for Evidence Synthesis. JBI. 10.46658/JBIMES-20-12

Piccolo, S.R., Frampton, M.B., 2016. Tools and techniques for computational reproducibility. GigaScience 5, 30. 10.1186/s13742-016-0135-4

Pitman, R., Fisman, D., Zaric, G.S., Postma, M., Kretzschmar, M., Edmunds, J., Brisson, M., 2012. Dynamic Transmission Modeling: A Report of the ISPOR-SMDM Modeling Good Research Practices Task Force-5. Value in Health 15, 828–834. 10.1016/j.jval.2012.06.011

Pokutnaya, D., Childers, B., Arcury-Quandt, A.E., Hochheiser, H., Panhuis, W.G.V., 2023a. An implementation framework to improve the transparency and reproducibility of computational models of infectious diseases. PLOS Computational Biology 19, e1010856. 10.1371/journal.pcbi.1010856

Pokutnaya, D., Van Panhuis, W.G., Childers, B., Hawkins, M.S., Arcury-Quandt, A.E., Matlack, M., Carpio, K., Hochheiser, H., 2023b. Inter-rater reliability of the infectious disease modeling reproducibility checklist (IDMRC) as applied to COVID-19 computational modeling research. BMC Infect Dis 23, 733. 10.1186/s12879-023-08729-4

Porgo, T.V., Norris, S.L., Salanti, G., Johnson, L.F., Simpson, J.A., Low, N., Egger, M., Althaus, C.L., 2019. The use of mathematical modeling studies for evidence synthesis and guideline development: A glossary. Res Synth Methods 10, 125–133. 10.1002/jrsm.1333

R Core Team, 2024. R: A Language and Environment for Statistical Computing. R Foundation for Statistical Computing, Vienna, Austria.

Ryckman, T., Luby, S., Owens, D.K., Bendavid, E., Goldhaber-Fiebert, J.D., 2020. Methods for Model Calibration under High Uncertainty: Modeling Cholera in Bangladesh. Med Decis Making 40, 693–709. 10.1177/0272989X20938683

Shaweno, D., Trauer, J.M., Denholm, J.T., McBryde, E.S., 2018. The role of geospatial hotspots in the spatial spread of tuberculosis in rural Ethiopia: a mathematical model. R Soc Open Sci 5, 180887. 10.1098/rsos.180887

Stout, N.K., Knudsen, A.B., Kong, C.Y., McMahon, P.M., Gazelle, G.S., 2009. Calibration Methods Used in Cancer Simulation Models and Suggested Reporting Guidelines. Pharmacoeconomics 27, 533–545. 10.2165/11314830-000000000-00000

Tricco, A.C., Lillie, E., Zarin, W., O’Brien, K.K., Colquhoun, H., Levac, D., Moher, D., Peters, M.D.J., Horsley, T., Weeks, L., Hempel, S., Akl, E.A., Chang, C., McGowan, J., Stewart, L., Hartling, L., Aldcroft, A., Wilson, M.G., Garritty, C., Lewin, S., Godfrey, C.M., Macdonald, M.T., Langlois, E.V., Soares-Weiser, K., Moriarty, J., Clifford, T., Tunçalp, Ö., Straus, S.E., 2018. PRISMA Extension for Scoping Reviews (PRISMA-ScR): Checklist and Explanation. Ann Intern Med 169, 467–473. 10.7326/M18-0850

Tuncer, N., Le, T.T., 2018. Structural and practical identifiability analysis of outbreak models. Mathematical biosciences 299, 1–18.

Unwin, H., Routledge, I., Flaxman, S., Rizoiu, M., Lai, S., Cohen, J., Weiss, D., Mishra, S., Bhatt, S., 2021. Using Hawkes Processes to model imported and local malaria cases in near-elimination settings. PLoS Computational Biology 17, 1–18. 10.1371/journal.pcbi.1008830

Vanni, T., Karnon, J., Madan, J., White, R.G., Edmunds, W.J., Foss, A.M., Legood, R., 2011. Calibrating models in economic evaluation: a seven-step approach. Pharmacoeconomics 29, 35–49. 10.2165/11584600-000000000-00000

Veritas Health Innovation, 2024. Covidence systematic review software.

Willem, L., Verelst, F., Bilcke, J., Hens, N., Beutels, P., 2017. Lessons from a decade of individual-based models for infectious disease transmission: a systematic review (2006-2015). BMC Infectious Diseases 17, 612. 10.1186/s12879-017-2699-8

