## Supplementary tables for "Calibration of transmission-dynamic infectious disease models: a scoping review and reporting framework"

**Table S1***: Inclusion and exclusion criteria based on the Studies, Data, Methods and Outcome (SDMO) framework.*

| SDMO | Element | Inclusion and exclusion criteria |
|---|---|---|
| **Studies** | Dynamic transmission models of HIV, TB or malaria in humans | Include:<br>• Published dynamic transmission modeling studies of HIV, TB or malaria in humans published from 1 January 2018 to 16 January 2024 (search date).<br><br>Exclude:<br>• Studies based on models that do not have a dynamic transmission component.<br>• Studies in which no model is proposed.<br>• Studies focused on diseases other than HIV, TB or malaria.<br>• Studies that do not involve human populations.<br>• Studies not available in the English language. |
| **Data** | Empirical data or published estimates. | Include:<br>• Studies which calibrate models to empirical data or published estimates.<br><br>Exclude:<br>• Studies which calibrate models to simulated data.<br>• Studies for which the data used is not mentioned or described. |
| **Method** | Model calibration | Include:<br>• Studies which calibrate their models to empirical data or published.<br>Exclude<br>• Studies that do not perform model calibration. |
| **Outcomes** | | Outcome measures in studies do not form part of the inclusion or exclusion criteria. |

***Table S2:*** *Preferred Reporting Items for Systematic reviews and Meta-Analyses extension for Scoping Reviews (PRISMA-ScR) Checklist*

| SECTION | ITEM | PRISMA-ScR CHECKLIST ITEM | REPORTED ON PAGE # |
|---|---|---|---|
| **TITLE** | | | |
| Title | 1 | Identify the report as a scoping review. | 1 |
| **ABSTRACT** | | | |
| Structured summary | 2 | Provide a structured summary that includes (as applicable): background, objectives, eligibility criteria, sources of evidence, charting methods, results, and conclusions that relate to the review questions and objectives. | 2 |
| **INTRODUCTION** | | | |
| Rationale | 3 | Describe the rationale for the review in the context of what is already known. Explain why the review questions/objectives lend themselves to a scoping review approach. | 5-6 |
| Objectives | 4 | Provide an explicit statement of the questions and objectives being addressed with reference to their key elements (e.g., population or participants, concepts, and context) or other relevant key elements used to conceptualize the review questions and/or objectives. | 5-6 |
| **METHODS** | | | |
| Protocol and registration | 5 | Indicate whether a review protocol exists; state if and where it can be accessed (e.g., a Web address); and if available, provide registration information, including the registration number. | 9 |
| Eligibility criteria | 6 | Specify characteristics of the sources of evidence used as eligibility criteria (e.g., years considered, language, and publication status), and provide a rationale. | 9-10 |
| Information sources* | 7 | Describe all information sources in the search (e.g., databases with dates of coverage and contact with authors to identify additional sources), as well as the date the most recent search was executed. | 10 |
| Search | 8 | Present the full electronic search strategy for at least 1 database, including any limits used, such that it could be repeated. | Table S3 |
| Selection of sources of evidence† | 9 | State the process for selecting sources of evidence (i.e., screening and eligibility) included in the scoping review. | 10-11 |
| Data charting process‡ | 10 | Describe the methods of charting data from the included sources of evidence (e.g., calibrated forms or forms that have been | 10-12 |

| SECTION | ITEM | PRISMA-ScR CHECKLIST ITEM | REPORTED ON PAGE # |
|---|---|---|---|
| | | tested by the team before their use, and whether data charting was done independently or in duplicate) and any processes for obtaining and confirming data from investigators. | |
| Data items | 11 | List and define all variables for which data were sought and any assumptions and simplifications made. | 11, data extraction form provided as supplementary material |
| Critical appraisal of individual sources of evidence§ | 12 | If done, provide a rationale for conducting a critical appraisal of included sources of evidence; describe the methods used and how this information was used in any data synthesis (if appropriate). | Not done or applicable to study |
| Synthesis of results | 13 | Describe the methods of handling and summarizing the data that were charted. | 11-13 |
| **RESULTS** | | | |
| Selection of sources of evidence | 14 | Give numbers of sources of evidence screened, assessed for eligibility, and included in the review, with reasons for exclusions at each stage, ideally using a flow diagram. | 13 |
| Characteristics of sources of evidence | 15 | For each source of evidence, present characteristics for which data were charted and provide the citations. | 13 |
| Critical appraisal within sources of evidence | 16 | If done, present data on critical appraisal of included sources of evidence (see item 12). | Not applicable to study |
| Results of individual sources of evidence | 17 | For each included source of evidence, present the relevant data that were charted that relate to the review questions and objectives. | Sheet S1 |
| Synthesis of results | 18 | Summarize and/or present the charting results as they relate to the review questions and objectives. | 14-18 |
| **DISCUSSION** | | | |
| Summary of evidence | 19 | Summarize the main results (including an overview of concepts, themes, and types of evidence available), link to the review questions and objectives, and consider the relevance to key groups. | 18-20 |
| Limitations | 20 | Discuss the limitations of the scoping review process. | 20-21 |
| Conclusions | 21 | Provide a general interpretation of the results with respect to the review questions and objectives, as well as potential implications and/or next steps. | 21-22 |

| SECTION | ITEM | PRISMA-ScR CHECKLIST ITEM | REPORTED ON PAGE # |
| --- | --- | --- | --- |
| **FUNDING** | | | |
| Funding | 22 | Describe sources of funding for the included sources of evidence, as well as sources of funding for the scoping review. Describe the role of the funders of the scoping review. | 23 |

JBI = Joanna Briggs Institute; PRISMA-ScR = Preferred Reporting Items for Systematic reviews and Meta-Analyses extension for Scoping Reviews.
* Where *sources of evidence* (see second footnote) are compiled from, such as bibliographic databases, social media platforms, and Web sites.
† A more inclusive/heterogeneous term used to account for the different types of evidence or data sources (e.g., quantitative and/or qualitative research, expert opinion, and policy documents) that may be eligible in a scoping review as opposed to only studies. This is not to be confused with *information sources* (see first footnote).
‡ The frameworks by Arksey and O'Malley (6) and Levac and colleagues (7) and the JBI guidance (4, 5) refer to the process of data extraction in a scoping review as data charting.
§ The process of systematically examining research evidence to assess its validity, results, and relevance before using it to inform a decision. This term is used for items 12 and 19 instead of "risk of bias" (which is more applicable to systematic reviews of interventions) to include and acknowledge the various sources of evidence that may be used in a scoping review (e.g., quantitative and/or qualitative research, expert opinion, and policy document).
*From:* Tricco AC, Lillie E, Zarin W, O'Brien KK, Colquhoun H, Levac D, et al. PRISMA Extension for Scoping Reviews (PRISMAScR): Checklist and Explanation. Ann Intern Med. 2018;169:467–473. doi: 10.7326/M18-0850.

**Table S3**: *Full electronic search* strategy*

| DATABASE (PLATFORM) | SEARCH | RESULTS |
|---|---|---|
| **PUBMED (NLM)** | (("Epidemiological Models"[Mesh] OR ("Transmission model"[tiab:~3] OR "transmission models"[tiab:~3] OR "dynamic model"[tiab:~3] OR "dynamic models"[tiab:~3] OR "mathematical model"[tiab:~3] OR "mathematical models"[tiab:~3] OR "simulation model"[tiab:~3] OR "Simulation models"[tiab:~3])) AND ("Malaria"[Mesh] OR malaria*[tiab] OR "plasmodium infection*"[tiab] OR "remittent fever*"[tiab] OR "Marsh fever*"[tiab] OR paludis*[tiab] OR "Tuberculosis"[Mesh] OR tuberculos*[tiab] OR tuberculoma*[tiab] OR koch-disease*[tiab] OR "HIV infections"[Mesh] OR "HIV"[Majr] OR HIV[tiab] OR "human immunodeficiency virus"[tiab] OR "Human immuno-deficiency virus"[tiab] OR HIVAIDS[tiab] OR "acquired immunodeficiency syndrome*"[tiab] OR "acquired immuno-deficiency syndrome*"[tiab])) NOT (“animals”[mesh] NOT “Humans”[mesh])<br>Date Range: from 1 January 2018 – 31 December 2023 | 1215 |
| **EMBASE (ELSEVIER)** | 1. ((‘Disease model’/exp/mj OR ‘Mathematical model’/exp/mj) AND ('disease transmission'/exp OR 'infection control'/exp)) OR ((transmission* OR dynamic* OR mathematical* OR simulation*) NEAR/3 model*):ti,ab,kw<br><br>2. 'Malaria'/exp OR 'Human immunodeficiency Virus'/exp/mj OR 'Human immunodeficiency virus infections'/exp OR 'Tuberculosis'/exp OR ('human immunodeficiency virus' OR HIV OR 'human immuno-deficiency virus' OR HIVAIDS OR 'acquired immunodeficiency sydrome*' OR 'acquired immuno-deficiency syndrome*' OR malaria* OR tuberculos* OR remittent-fever* OR marsh-fever* OR paludis* OR tuberculoma OR koch-disease*):ti,ab,kw<br><br>3. (('Nonhuman'/syn OR 'Animal'/syn) NOT 'Human'/exp)<br><br>4. #1 AND #2 NOT #3 AND [embase]/lim AND [2018-2023]/py AND ([article]/lim OR [article in press]/lim OR [data papers]/lim OR [review]/lim OR [preprint]/lim) | 1236 |
| **GLOBAL HEALTH (EBSCO)** | 1. DE (("disease models” OR "mathematical models" OR "simulation models") AND ("transmission" OR "infection control" OR "infectious diseases")) OR TI ((dynamic* OR transmission* OR mathematical* OR simulation*) N3 model*) OR AB ((dynamic* OR transmission* OR mathematical* OR simulation*) N3 model*)<br><br>2. DE ("malaria" OR "Blackwater fever" OR "Tuberculosis" OR "extrapulmonary tuberculosis" OR "miliary tuberculosis" OR "Human immunodeficiency viruses" OR "HIV Infections") OR TI ("human immunodeficiency virus" OR HIV OR "human | 1367 |

| DATABASE (PLATFORM) | SEARCH | RESULTS |
|---|---|---|
| | immuno-deficiency virus" OR HIVAIDS OR "acquired immunodeficiency sydrome*" OR "acquired immuno-deficiency syndrome*" OR malaria* OR tuberculos* OR remittent-fever* OR marsh-fever* OR paludis* OR tuberculoma OR koch-disease*) OR AB ("human immunodeficiency virus" OR HIV OR "human immuno-deficiency virus" OR HIVAIDS OR "acquired immunodeficiency sydrome*" OR "acquired immuno-deficiency syndrome*" OR malaria* OR tuberculos* OR remittent-fever* OR marsh-fever* OR paludis* OR tuberculoma OR koch-disease*)<br><br>S1 AND S2<br>Limiters - Publication Year: 20180101-20231231 | |
| **WEB OF SCIENCE CORE COLLECTION – SCI-EXP & ESCI (CLARIVATE)** | #1 (TI=(model* NEAR/3 (mathematical* OR simulation$ OR transmission OR dynamic$))) OR (AB=(model* NEAR/3 (mathematical* OR simulation$ OR transmission OR dynamic$))) OR (AK=(model* NEAR/3 (mathematical OR simulation$ OR transmission OR dynamic$)))<br><br>#2 (TI=("human immunodeficiency virus" OR HIV OR "human immuno-deficiency virus" OR HIVAIDS OR "acquired immunodeficiency sydrome*" OR "acquired immuno-deficiency syndrome*" OR malaria* OR tuberculos* OR remittent-fever* OR marsh-fever* OR paludis* OR tuberculoma OR koch-disease*)) OR (AB=("human immunodeficiency virus" OR HIV OR "human immuno-deficiency virus" OR HIVAIDS OR "acquired immunodeficiency sydrome*" OR "acquired immuno-deficiency syndrome*" OR malaria* OR tuberculos* OR remittent-fever* OR marsh-fever* OR paludis* OR tuberculoma OR koch-disease*)) OR (AK=("human immunodeficiency virus" OR HIV OR "human immuno-deficiency virus" OR HIVAIDS OR "acquired immunodeficiency sydrome*" OR "acquired immuno-deficiency syndrome*" OR malaria* OR tuberculos* OR remittent-fever* OR marsh-fever* OR paludis* OR tuberculoma OR koch-disease*))<br><br>#1 AND #2 AND 2018-2023 | 2359 |
| **GLOBAL INDEX MEDICUS – ALL INDICES (WHO)** | tw:((transmission OR dynamic OR simulation OR mathematical) AND model*) AND tw:("human immunodeficiency virus" OR HIV OR "human immuno-deficiency virus" OR HIVAIDS OR "acquired immunodeficiency sydrome*" OR "acquired immuno-deficiency syndrome*" OR malaria* OR tuberculos* OR remittent-fever* OR marsh-fever* OR paludis* OR tuberculoma OR koch-disease*)<br>Date limit: 2018-2023 | 55 |
| **FROM WEBSITES** | | 286 |

| DATABASE (PLATFORM) | SEARCH | RESULTS |
|---|---|---|
| **TOTAL** | | 6518 |
| **AFTER DEDUPLICATION** | | 3138 |

Grey Literature Search

| WEBSITE URL | # OF ARTICLES RETRIEVED |
|---|---|
| **HTTP://HIVMODELING.ORG/PUBLICATIONS-REPORTS** | 21 |
| **TB-MAC NEWSLETTERS** | 265 |
| **TOTAL** | 286 |

*Searches run on 16 January 2024 by Emmanuelle A Dankwa

**Table S4:** *Details of studies with multiple calibrated models.*

| STUDY DOI | FULL TITLE | NO. CALIBRATED MODELS |
|---|---|---|
| https:// doi.org/10.1371/journal.pone.0242595 | Challenges in estimating HIV prevalence trends and geographical variation in HIV prevalence using antenatal data: Insights from mathematical modelling | 2 |
| https://doi.org/10.1097/ede.0000000000001418 | The Health and Economic Benefits of Tests That Predict Future Progression to Tuberculosis Disease | 3 |
| http://doi.org/10.1097/qad.0000000000002826 | Mathematical modelling of the influence of serosorting on the population-level HIV transmission impact of pre-exposure prophylaxis | 2 |
| https://doi.org/10.1164/rccm.201907-1289oc | Comparative Modeling of Tuberculosis Epidemiology and Policy Outcomes in California. | 3 |
| https://doi.org/10.1371/journal.pone.0199453 | The emerging health impact of voluntary medical male circumcision in Zimbabwe: An evaluation using three epidemiological models. | 3 |

**Table S5**: Description of calibration methods used in models. For each method, we provide up to three references in our review demonstrating applications of the method. Where fewer than three articles use a method, all articles using that method are listed.

| # | Method | Short description | Examples of articles in review applying method |
|---|---|---|---|
| 1 | Markov Chain Monte Carlo (MCMC) | Markov Chain Monte Carlo (MCMC) is a class of algorithms used to sample from probability distributions that are complex or high-dimensional, making them difficult to analyze using traditional methods. A Markov chain is a stochastic process where the future state depends only on the current state, not on the sequence of events that preceded it. This property is known as the Markov property. In MCMC, this concept is used to generate a sequence of samples that approximate the target distribution. See (van de Schoot et al., 2021) for a primer. | (Arinaminpathy et al., 2023; Nsengiyumva et al., 2022; Xue et al., 2022)09/03/2025 16:34:00 |
| 2 | Approximate Bayesian Computation (ABC) | Approximate Bayesian Computation (ABC) (Beaumont et al., 2002) is a class of computational methods in Bayesian statistics that allows for parameter estimation and model selection when the likelihood function is intractable or too expensive to evaluate. It does not require explicitly evaluating a likelihood function, uses a distance function to measure the discrepancy between simulated and observed data, and has a tolerance parameter to threshold how close simulated data must be to the observed data. | (Fraser et al., 2021; Horton et al., 2022; Sahu et al., 2023) |
| 3 | Incremental Mixture Importance Sampling (IMIS) | Incremental Mixture Importance Sampling (IMIS) (Raftery and Bao, 2010) is an advanced statistical technique used for efficient sampling from complex probability distributions. IMIS involves drawing samples from a prior distribution, calculating importance weights based on a target distribution, identifying regions of high importance, adding new mixture components to high weight regions, and then repeating these steps until a stopping criterion is met. Final samples are generated by resampling from an accumulated mixture distribution. | (Chang et al., 2018; Menzies et al., 2021; Yang et al., 2021) |
| 4 | Sequential Monte Carlo (SMC) | SMC methods (Doucet et al., 2001) or particle filtering methods aim to approximate posterior distributions by using a set of weighted samples (particles) that are sequentially updated as new data becomes available. Unlike Monte Carlo methods, SMC processes data sequentially. It involves the concept of importance sampling, which involves drawing samples from a proposal distribution and weighting them to approximate a target distribution. | (Lima et al., 2021; Rodó et al., 2021; Vliet et al., 2019)09/03/2025 16:34:00 |

| # | Method | Short description | Examples of articles in review applying method |
|---|---|---|---|
| 5 | Sampling importance resampling (SIR) | The sampling importance resampling (SIR) (Skare et al., 2003) algorithm's goal is to draw a random sample at a target distribution. The algorithm works by drawing an independent random sample from a proposal distribution, within which a second smaller sample is drawn (with or without replacement) with a probability. Ultimately, this method complains importance sampling with resampling to improve sample quality. | (Doyle et al., 2022; Salvatore et al., 2019; Silhol et al., 2021) |
| 6 | History matching with emulation (HME) | History matching (Iskauskas et al., 2023; Scarponi et al., 2023b) is an iterative process that involves exploring the parameter space of a model to find non-implausible parameter sets that could result in outputs consistent with observed data. This process is conducted in waves to eliminate implausible regions of parameter space. Emulation is performed using emulators that are meant to approximate the simulated output with a degree of uncertainty. | (McCreesh et al., 2018; McGillen et al., 2018; Scarponi et al., 2023a) |
| 7 | Laplace Approximation | The Laplace approximation (Kristensen et al., 2016) is a tool that provides a Gaussian approximation to a continuous probability density function. Often in model calibration it is used to provide an approximation of a posterior distribution of model parameters. | (Maheu-Giroux et al., 2019) |
| 8 | Least squares estimation | Least squares estimation (van de Geer, 2005) operates by attempting to minimize the sum of squares of residuals comparing observed values to model output values. The parameters that minimize this function are the most likely parameters in the constructed model. | (Awine and Silal, 2022; Ochieng, 2024; Torres et al., 2023) |
| 9 | Maximum likelihood estimation | Maximum Likelihood Estimation (MLE) (Myung, 2003) involves finding the parameter values that maximize the likelihood of observing real-world data, which serves as the calibration target. The likelihood function quantifies the probability of this observed data given a set of model parameters, and the MLE parameters are those that optimize this function. | (Hauser et al., 2019; Mettle et al., 2020; Smith et al., 2022) |
| 10 | Manual (hand-tuning) | Manual (hand-tuning) involves adjusting model parameters by trial and error to closely align model predictions with observed (i.e. calibration target) data. For instance, the accuracy of model predictions is often assessed through visual comparison, by plotting them against observed data over time to check if trends align. Alternatively, model outputs can be evaluated against confidence intervals | (Cilloni et al., 2020; Estill et al., 2023; Modu et al., 2023) |

| # | Method | Short description | Examples of articles in review applying method |
| --- | --- | --- | --- |
| | | around observed data, providing a range-based assessment of fit that shows whether predictions fall within an acceptable error margin. | |
| 11 | Nelder-Mead algorithm | The Nelder-Mead algorithm (Nelder and Mead, 1965) is an optimization method used to find parameter values that minimize the difference between model predictions and observed (i.e. calibration target) data, optimizing the goodness-of-fit (GoF) metric—often the mean percentage deviation across studies included in a review. It is a simplex-based approach that avoids using gradients or derivatives; instead, it evaluates the objective function at multiple points (forming a 'simplex') and iteratively moves toward better values. This heuristic method is especially useful for models where derivatives are challenging to compute or where the objective function is not smooth. | (Hsieh et al., 2020; Krebs et al., 2019; Zang et al., 2020) |
| 12 | Grey Wolf Optimizer (GWO) | Guo et al. (2021) explain: "The GWO algorithm mimics the leadership hierarchy and hunting mechanism of grey wolves in nature. Four types of grey wolves such as alpha, beta, delta, and omega are employed for simulating the leadership hierarchy. In addition, the three main steps of hunting, searching for prey, encircling prey, and attacking prey, are implemented." In the context of the Grey Wolf Optimizer (GWO), the 'wolves' represent candidate solutions in the search space of the optimization problem, with each wolf corresponding to a set of parameter values in the model being optimized. During the optimization process, each wolf's position (the set of parameter values it represents) is adjusted through three main phases: searching (exploring the search space by randomly adjusting positions), encircling (refining positions around the prey by narrowing the search space), and attacking (aggressively refining positions by exploiting the most promising solutions to focus on local optimization and approach the optimal solution). The 'prey' in this context refers to the optimal solution. See Mirjalili et al. (2014) for details. | (Guo and Zhang, 2023; Guo et al., 2023, 2021) |
| 13 | Genetic algorithm | Based on the process of natural selection, the genetic algorithm (Lucasius and Kateman, 1993) is iterative with the following general steps: at each iteration, 1) select a random population of candidate solutions (“individuals”) each with specific characteristic; 2) evaluate the suitability (“fitness”) of each candidate solution based on the value of the objective function, 3) candidates with better values for the objective function are randomly selected from the current set of candidates, 4) | (Chen et al., 2023; de Oliveira et al., 2022) |

| # | Method | Short description | Examples of articles in review applying method |
|---|---|---|---|
| | | selected candidates are modified ("recombined" or "mutated") to produce a new set of candidates. This new set of candidates are used in the next iteration and steps 1)-5) are repeated for this set. The result of each iteration is referred to as a "generation" and the algorithm terminates when either a defined number of generations have been produced or when candidates meet certain defined requirements for "fitness". Genetic algorithms belong to a wider class of *evolutionary algorithms* (Vikhar, 2016). | |
| 14 | Parallel Simultaneous perturbation Optimisation (PSPO) | Parallel Simultaneous Perturbation Optimization (PSPO) (Alaeddini and Klein, 2019) works by simultaneously perturbing multiple parameters and evaluating the resulting changes in the objective function, typically the error between predicted and target data. This method is computationally efficient, as it requires fewer model evaluations compared to traditional gradient-based optimization techniques. By performing parallel evaluations, PSPO accelerates the calibration process while maintaining accuracy in parameter estimation. | (Akullian et al., 2020; Bershteyn et al., 2018) |
| 15 | Broyden-Fletcher-Goldfarb-Shanno algorithm | The Broyden-Fletcher-Goldfarb-Shanno (BFGS) algorithm (Fletcher, 1987) is an iterative optimization method that belongs to a class of second-order algorithms called Quasi-Newton methods. These algorithms approximate the second derivative (the Hessian matrix) of an objective function, which helps determine the optimal direction for adjusting model parameters. Since exact second derivatives are often difficult or costly to compute, BFGS uses gradient evaluations to iteratively improve its approximation of the Hessian. It is commonly used to optimize complex objective functions, such as those in infectious disease modeling, where exact second derivatives are difficult or costly to compute. | (Doyle et al., 2022; Maheu-Giroux et al., 2019) |
| 16 | Coordinate descent algorithm | Coordinate descent algorithms (Wright, 2015) work iteratively by fixing, at each iteration,the majority of current values of the parameters being optimized, then minimizing the objective function with respect to the remaining values. Consequently, each minimization problem has a lower dimension than the previous problem and is therefore easier to solve. | (Guo et al., 2020) |
| 17 | Deviance-based loss | As used in (Wang et al., 2022), the deviance-based loss method minimizes a deviance score: a statistic which compares a model of interest to the *saturated* | (Wang et al., 2022) |

| # | Method | Short description | Examples of articles in review applying method |
|---|---|---|---|
| | | (perfect fit) model using the maximum likelihood estimates of a parameter (set) under each model. | |
| 18 | Levenberg-Marquardt optimization algorithm | The Levenberg-Marquardt (LM) algorithm (Gavin, 2024; Levenberg, 1944; Marquardt, 1963) is an iterative algorithm for nonlinear least squares problems: minimization problems in which the parameters are related to model predicted in a nonlinear fashion which is typically the case for many disease models. The LM algorithm combines gradient descent and Gauss-Newton methods (Wang, 2012) to reach optimal parameter values. The gradient descent method minimizes an objective function by updating coefficient values in the direction of the steepest descent of the objective function, which is the direction opposite to its gradient. | (Arregui et al., 2018) |
| 19 | Local sequential quadratic programming (DESQP) optimization algorithm | Wu et al. (2020) explain: "DESQP, which combines differential evolution (DE) and local sequential quadratic programming (SQP), is a method used to search for the optimal solution of DE. In the method, DE is used as a base level search and SQP is used as a local search. DE is first applied to the short term of the problem to find the best solution. This optimal solution is given to SQP as an initial condition to fine tune the solution to reach the global optimum or near global optimum." | (Wu et al., 2020) |
| 20 | Maximum-a-Posteriori (MAP) estimation | MAP estimation, based on Bayes theorem, involves estimating the mode of a posterior density. In performing MAP estimation, one maximizes the posterior probability of the parameters $\theta$ given the data set $X,$ given by $P(X\|\theta) * P(\theta)$, over a range of $\theta$ values. MAP estimation is equivalent to the maximum likelihood estimation under the assumption of a uniform prior: where all $\theta$ values are equally likely. | (Routledge et al., 2020) |
| 21 | Mean square error with threshold | The mean square error estimation method is based on minimizing the mean square error: the squared distance between the observed data and the simulated data. | (Zwick et al., 2021) |
| 22 | Method of moments | As used in (Adeyemo et al., 2023), the method of moments involves: (1) expressing population moments (in this case, the mean) as a function of parameters to be estimated, (2) equating expression from (1) to the sample mean, (3) solving equations to obtain solutions for parameters. | (Adeyemo et al., 2023) |

| # | Method | Short description | Examples of articles in review applying method |
|---|---|---|---|
| 23 | Minimum Step Deviation | See "Model Calibration" section in Supplementary material of (Tan et al., 2019). | (Tan et al., 2019) |
| 24 | Minimum contrast estimation | Minimum contrast estimation is an inference method for diffusion models and suitable for cases in which transition probability densities are unavailable in closed form hence an approximation of the (log) likelihood is needed. This process is explained in detail and applied in (Abou-Bakre and El Maroufy, 2018). | (Abou-Bakre and El Maroufy, 2018) |
| 25 | Root Mean Squared (RMS) deviation | The root mean squared (RMS) deviation method is based on minimizing a RMS objective function: the square root of the average squared residuals, which is the difference between the model prediction and the observed data (target). | (N'Diaye et al., 2019) |

### References for Table S5


Abou-Bakre, A., El Maroufy, H., 2018. Parameter inference for HIV stochastic diffusion model in closed heterosexual population. Math. Methods Appl. Sci. 41, 9081–9091. https://doi.org/10.1002/mma.4940

Adeyemo, S., Sangotola, A., Korosteleva, O., 2023. Modeling Transmission Dynamics of Tuberculosis–HIV Co-Infection in South Africa. Epidemiologia 4, 408–419. https://doi.org/10.3390/epidemiologia4040036

Akullian, A., Morrison, M., Garnett, G.P., Mnisi, Z., Lukhele, N., Bridenbecker, D., Bershteyn, A., 2020. The effect of 90-90-90 on HIV-1 incidence and mortality in eSwatini: a mathematical modelling study. Lancet HIV 7, e348. https://doi.org/10.1016/S2352-3018(19)30436-9

Alaeddini, A., Klein, D.J., 2019. Parallel Simultaneous Perturbation Optimization. Asia-Pac. J. Oper. Res. 36, 1950009. https://doi.org/10.1142/S021759591950009X

Arinaminpathy, N., Rade, K., Kumar, R., Joshi, R.P., Rao, R., 2023. The potential impact of vaccination on tuberculosis burden in India: A modelling analysis. Indian J. Med. Res. 157, 119. https://doi.org/10.4103/ijmr.ijmr_328_23

Arregui, S., Iglesias, M.J., Samper, S., Marinova, D., Martin, C., Sanz, J., Moreno, Y., 2018. Data-driven model for the assessment of Mycobacterium tuberculosis transmission in evolving demographic structures. Proc. Natl. Acad. Sci. 115, E3238–E3245. https://doi.org/10.1073/pnas.1720606115

Awine, T., Silal, S.P., 2022. Assessing the effectiveness of malaria interventions at the regional level in Ghana using a mathematical modelling application. PLOS Glob. Public Health 2, e0000474. https://doi.org/10.1371/journal.pgph.0000474

Beaumont, M.A., Zhang, W., Balding, D.J., 2002. Approximate Bayesian computation in population genetics. Genetics 162, 2025–2035.

Bershteyn, A., Mutai, K.K., Akullian, A.N., Klein, D.J., Jewell, B.L., Mwalili, S.M., 2018. The influence of mobility among high-risk populations on HIV transmission in Western Kenya. Infect. Dis. Model. 3, 97–106. https://doi.org/10.1016/j.idm.2018.04.001

Chang, S.T., Chihota, V.N., Fielding, K.L., Grant, A.D., Houben, R.M., White, R.G., Churchyard, G.J., Eckhoff, P.A., Wagner, B.G., 2018. Small contribution of gold mines to the ongoing tuberculosis epidemic in South Africa: a modeling-based study. BMC Med. 16, 52. https://doi.org/10.1186/s12916-018-1037-3

Chen, N., Chen, S., Li, X., Li, Z., Chen, N., Chen, S., Li, X., Li, Z., 2023. Modelling and analysis of the HIV/AIDS epidemic with fast and slow asymptomatic infections in China from 2008 to 2021. Math. Biosci. Eng. 20, 20770–20794. https://doi.org/10.3934/mbe.2023919

Cilloni, L., Kranzer, K., Stagg, H.R., Arinaminpathy, N., 2020. Trade-offs between cost and accuracy in active case finding for tuberculosis: A dynamic modelling analysis. PLoS Med. 17, e1003456. https://doi.org/10.1371/journal.pmed.1003456

de Oliveira, R.B., Rubio, F.A., Anderle, R., Sanchez, M., de Souza, L.E., Macinko, J., Dourado, I., Rasella, D., 2022. Incorporating social determinants of health into the mathematical modeling of HIV/AIDS. Sci. Rep. 12, 20541. https://doi.org/10.1038/s41598-022-24459-0

Doucet, A., de Freitas, N., Gordon, N., 2001. Sequential Monte Carlo methods in practice, Statistics for engineering and information science. Springer, New York.

Doyle, C., Cox, J., Milwid, R., Bitera, R., Delaunay, C., Alary, M., Lambert, G., Tremblay, C., Mishra, S., Maheu-Giroux, M., 2022. Measuring progress towards reaching zero new HIV acquisitions among key populations in Québec (Canada) using routine surveillance data: a mathematical modelling study. J. Int. AIDS Soc. 25, e25994. https://doi.org/10.1002/jia2.25994

Estill, J., Xun, Y., Wu, S., Hu, L., Yang, N., Yang, S., Chen, Y., Li, G., 2023. Tuberculosis screening among children and adolescents in China: insights from a mathematical model. Intell. Med. 3, 157–163. https://doi.org/10.1016/j.imed.2022.09.001

Fletcher, R. (Roger), 1987. Practical methods of optimization. Chichester ; New York : Wiley.

Fraser, H., Borquez, A., Stone, J., Abramovitz, D., Brouwer, K.C., Goodman-Meza, D., Hickman, M., Patterson, T.L., Silverman, J., Smith, L., Strathdee, S.A., Martin, N.K., Vickerman, P., 2021. Overlapping Key Populations and HIV Transmission in Tijuana, Mexico: A Modelling Analysis of Epidemic Drivers. AIDS Behav. 25, 3814–3827. https://doi.org/10.1007/s10461-021-03361-2

Gavin, H.P., 2024. The Levenberg-Marquardt algorithm for nonlinear least squares curve-fitting problems.

Guo, Z., Xiao, D., Xu, S., He, K., 2020. Analysis and forecast of the HIV/AIDS epidemic in Mainland China, 1985-2016. J. Public Health Oxf. Engl. 42, E458–E467. https://doi.org/10.1093/pubmed/fdz116

Guo, Z., Zhang, L., 2023. Global Dynamics of an Age-Structured Tuberculosis Model with Vaccine Failure and Nonlinear Infection Force. Axioms 12, 805. https://doi.org/10.3390/axioms12090805

Guo, Z.-K., Huo, H.-F., Xiang, H., Ren, Q.-Y., 2023. Global dynamics of a tuberculosis model with age-dependent latency and time delays in treatment. J. Math. Biol. 87, 66. https://doi.org/10.1007/s00285-023-01999-1

Guo, Z.-K., Xiang, H., Huo, H.-F., 2021. Analysis of an age-structured tuberculosis model with treatment and relapse. J. Math. Biol. 82, 45. https://doi.org/10.1007/s00285-021-01595-1

Hauser, A., Kusejko, K., Johnson, L.F., Wandeler, G., Riou, J., Goldstein, F., Egger, M., Kouyos, R.D., 2019. Bridging the gap between HIV epidemiology and antiretroviral resistance evolution: Modelling the spread of resistance in South Africa. PLoS Comput. Biol. 15, e1007083. https://doi.org/10.1371/journal.pcbi.1007083

Horton, K.C., White, R.G., Hoa, N.B., Nguyen, H.V., Bakker, R., Sumner, T., Corbett, E.L., Houben, R.M.G.J., 2022. Population benefits of addressing programmatic and social determinants of gender disparities in tuberculosis in Viet Nam: A modelling study. PLOS Glob. Public Health 2, e0000784. https://doi.org/10.1371/journal.pgph.0000784

Hsieh, Y., Jahn, A., Menzies, N., Yaesoubi, R., Salomon, J., Girma, B., Gunde, L., Eaton, J., Auld, A., Odo, M., Kiyiika, C., Kalua, T., Chiwandira, B., Mpunga, J., Mbendra, K., Corbett, L., Hosseinipour, M., Cohen, T., Kunkel, A., 2020. Evaluation of 6-Month Versus Continuous Isoniazid Preventive Therapy for Mycobacterium tuberculosis in Adults Living With HIV/AIDS in Malawi. JAIDS-J. Acquir. IMMUNE Defic. Syndr. 85, 643–650. https://doi.org/10.1097/QAI.0000000000002497

Iskauskas, A., Vernon, I., Goldstein, M., Scarponi, D., McKinley, T.J., White, R.G., McCreesh, N., 2023. Emulation and History Matching using the hmer Package. https://doi.org/10.48550/arXiv.2209.05265

Krebs, E., Enns, B., Wang, L., Zang, X., Panagiotoglou, D., Del Rio, C., Dombrowski, J., Feaster, D.J., Golden, M., Granich, R., Marshall, B., Mehta, S.H., Metsch, L., Schackman, B.R., Strathdee, S.A., Nosyk, B., localized HIV modeling study group, 2019. Developing a dynamic HIV transmission model for 6 U.S. cities: An evidence synthesis. PloS One 14, e0217559. https://doi.org/10.1371/journal.pone.0217559

Kristensen, K., Nielsen, A., Berg, C.W., Skaug, H., Bell, B.M., 2016. TMB: Automatic Differentiation and Laplace Approximation. J. Stat. Softw. 70, 1–21. https://doi.org/10.18637/jss.v070.i05

Levenberg, K., 1944. A method for the solution of certain non-linear problems in least squares. Q. Appl. Math. 2, 164–168. https://doi.org/10.1090/qam/10666

Lima, V.D., Zhu, J., Card, K.G., Lachowsky, N.J., Chowell-Puente, G., Wu, Z., Montaner, J.S.G., 2021. Can the combination of TasP and PrEP eliminate HIV among MSM in British Columbia, Canada? Epidemics 35, 100461. https://doi.org/10.1016/j.epidem.2021.100461

Lucasius, C.B., Kateman, G., 1993. Understanding and using genetic algorithms Part 1. Concepts, properties and context. Chemom. Intell. Lab. Syst. 19, 1–33. https://doi.org/10.1016/0169-7439(93)80079-W

Maheu-Giroux, M., Marsh, K., Doyle, C.M., Godin, A., Lanièce Delaunay, C., Johnson, L.F., Jahn, A., Abo, K., Mbofana, F., Boily, M.-C., Buckeridge, D.L., Hankins, C.A., Eaton, J.W., 2019. National HIV testing and diagnosis coverage in sub-Saharan Africa: a new modeling tool for estimating the “first 90” from program and survey data. AIDS Lond. Engl. 33 Suppl 3, S255–S269. https://doi.org/10.1097/QAD.0000000000002386

Marquardt, D.W., 1963. An Algorithm for Least-Squares Estimation of Nonlinear Parameters. J. Soc. Ind. Appl. Math. 11, 431–441.

McCreesh, N., Andrianakis, I., Nsubuga, R., Strong, M., Vernon, I., McKinley, T., Oakley, J., Goldstein, M., Hayes, R., White, R., 2018. Choice of time horizon critical in estimating costs and effects of changes to HIV programmes. PLOS ONE 13, e0196480. https://doi.org/10.1371/journal.pone.0196480

McGillen, J., Stover, J., Klein, D., Xaba, S., Ncube, G., Mhangara, M., Chipendo, G., Taramusi, I., Beacroft, L., Hallett, T., Odawo, P., Manzou, R., Korenromp, E., 2018. The emerging health impact of voluntary medical male circumcision in Zimbabwe: An evaluation using three epidemiological models. PLOS ONE 13, e0199453. https://doi.org/10.1371/journal.pone.0199453

Menzies, N.A., Swartwood, N., Testa, C., Malyuta, Y., Hill, A.N., Marks, S.M., Cohen, T., Salomon, J.A., 2021. Time Since Infection and Risks of Future Disease for Individuals with Mycobacterium tuberculosis Infection in the United States. Epidemiol. Camb. Mass 32, 70–78. https://doi.org/10.1097/EDE.0000000000001271

Mettle, F.O., Osei Affi, P., Twumasi, C., 2020. Modelling the Transmission Dynamics of Tuberculosis in the Ashanti Region of Ghana. Interdiscip. Perspect. Infect. Dis. 2020, 4513854. https://doi.org/10.1155/2020/4513854

Mirjalili, S., Mirjalili, S.M., Lewis, A., 2014. Grey Wolf Optimizer. Adv. Eng. Softw. 69, 46–61. https://doi.org/10.1016/j.advengsoft.2013.12.007

Modu, B., Polovina, N., Konur, S., 2023. Agent-Based Modeling of Malaria Transmission. IEEE ACCESS 11, 19794–19808. https://doi.org/10.1109/ACCESS.2023.3248292

Myung, I.J., 2003. Tutorial on maximum likelihood estimation. J. Math. Psychol. 47, 90–100. https://doi.org/10.1016/S0022-2496(02)00028-7

N'Diaye, D.S., Nsengiyumva, N.P., Uppal, A., Oxlade, O., Alvarez, G.G., Schwartzman, K., 2019. The potential impact and cost-effectiveness of tobacco reduction strategies for tuberculosis prevention in Canadian Inuit communities. BMC Med. 17, 26. https://doi.org/10.1186/s12916-019-1261-5

Nelder, J.A., Mead, R., 1965. A Simplex Method for Function Minimization. Comput. J. 7, 308–313. https://doi.org/10.1093/comjnl/7.4.308

Nsengiyumva, N.P., Campbell, J.R., Oxlade, O., Vesga, J.F., Lienhardt, C., Trajman, A., Falzon, D., Boon, S.D., Arinaminpathy, N., Schwartzman, K., 2022. Scaling up target regimens for tuberculosis preventive treatment in Brazil and South Africa: An analysis of costs and cost-effectiveness. PLOS Med. 19, e1004032. https://doi.org/10.1371/journal.pmed.1004032

Ochieng, F.O., 2024. SEIRS model for malaria transmission dynamics incorporating seasonality and awareness campaign. Infect. Dis. Model. 9, 84–102. https://doi.org/10.1016/j.idm.2023.11.010

Raftery, A.E., Bao, L., 2010. Estimating and Projecting Trends in HIV/AIDS Generalized Epidemics Using Incremental Mixture Importance Sampling. Biometrics 66, 1162–1173. https://doi.org/10.1111/j.1541-0420.2010.01399.x

Rodó, X., Martinez, P., Siraj, A., Pascual, M., 2021. Malaria trends in Ethiopian highlands track the 2000 "slowdown" in global warming. Nat. Commun. 12, 1555. https://doi.org/10.1038/s41467-021-21815-y

Routledge, I., Lai, S., Battle, K., Ghani, A., Gomez-Rodriguez, M., Gustafson, K., Mishra, S., Unwin, J., Proctor, J., Tatem, A., Li, Z., Bhatt, S., 2020. Tracking progress towards malaria elimination in China: Individual-level estimates of transmission and its spatiotemporal variation using a diffusion network approach. PLOS Comput. Biol. 16, e1007707. https://doi.org/10.1371/journal.pcbi.1007707

Sahu, M., Bayer, C.J., Roberts, D.A., Rooyen, H. van, Heerden, A. van, Shahmanesh, M., Asiimwe, S., Sausi, K., Sithole, N., Ying, R., Rao, D.W., Krows, M.L., Shapiro, A.E., Baeten, J.M., Celum, C., Revill, P., Barnabas, R.V., 2023. Population health impact, cost-effectiveness, and affordability of community-based HIV treatment and monitoring in South Africa: A health economics modelling study. PLOS Glob. Public Health 3, e0000610. https://doi.org/10.1371/journal.pgph.0000610

Salvatore, P.P., Kendall, E.A., Seabrook, D., Brown, J., Durham, G.H., Dowdy, D.W., 2019. Projecting the impact of variable MDR-TB transmission efficiency on long-term epidemic trends in South Africa and Vietnam. Sci. Rep. 9, 18099. https://doi.org/10.1038/s41598-019-54561-9

Scarponi, D., Clark, R.A., Weerasuriya, C.K., Emery, J., Houben, R.M.G.J., White, R., McCreesh, N., 2023a. Is neglect of self-clearance biasing TB vaccine impact estimates? BMJ Glob. Health 8, e012799. https://doi.org/10.1136/bmjgh-2023-012799

Scarponi, D., Iskauskas, A., Clark, R.A., Vernon, I., McKinley, T.J., Goldstein, M., Mukandavire, C., Deol, A., Weerasuriya, C., Bakker, R., White, R.G., McCreesh, N., 2023b. Demonstrating multi-country calibration of a tuberculosis model using new history matching and emulation package - *hmer*. Epidemics 43, 100678. https://doi.org/10.1016/j.epidem.2023.100678

Silhol, R., Geidelberg, L., Mitchell, K., Mishra, S., Dimitrov, D., Bowring, A., Béhanzin, L., Guédou, F., Diabaté, S., Schwartz, S., Billong, S., Njindam, I., Levitt, D., Mukandavire, C., Maheu-Giroux, M., Rönn, M., Dalal, S., Vickerman, P., Baral, S., Alary, M., Boily, M., 2021. Assessing the Potential Impact of Disruptions Due to COVID-19 on HIV Among Key and Lower-Risk Populations in the Largest Cities of Cameroon and Benin. JAIDS J. Acquir. Immune Defic. Syndr. 87, 899–911. https://doi.org/10.1097/QAI.0000000000002663

Skare, Ø., Bølviken, E., Holden, L., 2003. Improved Sampling-Importance Resampling and Reduced Bias Importance Sampling. Scand. J. Stat. 30, 719–737. https://doi.org/10.1111/1467-9469.00360

Smith, J., Oeltmann, J., Hill, A., Tobias, J., Boyd, R., Click, E., Finlay, A., Mondongo, C., Zetola, N., Moonan, P., 2022. Characterizing tuberculosis transmission dynamics in high-burden urban and rural settings. Sci. Rep. 12, 1–12. https://doi.org/10.1038/s41598-022-10488-2

Tan, J., Altice, F.L., Madden, L.M., Zelenev, A., 2019. The Impact of Expanding Opioid Agonist Therapies on HIV epidemic and mortality in Ukraine: a Modeling study. Lancet HIV 7, e121. https://doi.org/10.1016/S2352-3018(19)30373-X

Torres, M., Tubay, J., de losReyes, A., 2023. Quantitative Assessment of a Dual Epidemic Caused by Tuberculosis and HIV in the Philippines. Bull. Math. Biol. 85, 56. https://doi.org/10.1007/s11538-023-01156-1

van de Geer, S.A., 2005. Least Squares Estimation, in: Encyclopedia of Statistics in Behavioral Science. John Wiley & Sons, Ltd. https://doi.org/10.1002/0470013192.bsa199

van de Schoot, R., Depaoli, S., King, R., Kramer, B., Märtens, K., Tadesse, M.G., Vannucci, M., Gelman, A., Veen, D., Willemsen, J., Yau, C., 2021. Bayesian statistics and modelling. Nat. Rev. Methods Primer 1, 1–26. https://doi.org/10.1038/s43586-020-00001-2

Vikhar, P.A., 2016. Evolutionary algorithms: A critical review and its future prospects, in: 2016 International Conference on Global Trends in Signal Processing, Information Computing and Communication (ICGTSPICC). Presented at the 2016 International Conference on Global Trends in Signal Processing, Information Computing and Communication (ICGTSPICC), pp. 261–265. https://doi.org/10.1109/ICGTSPICC.2016.7955308

Vliet, M.M., Hendrickson, C., Nichols, B.E., Boucher, C.A., Peters, R.P., Vijver, D.A., 2019. Epidemiological impact and cost-effectiveness of providing long-acting pre-exposure prophylaxis to injectable contraceptive users for HIV prevention in South Africa: a modelling study. J. Int. AIDS Soc. 22, N.PAG-N.PAG.

Wang, Y., 2012. Gauss–Newton method. WIREs Comput. Stat. 4, 415–420. https://doi.org/10.1002/wics.1202

Wang, Y., Tanuma, J., Li, J., Iwahashi, K., Peng, L., Chen, C., Hao, Y., Gilmour, S., 2022. Elimination of HIV transmission in Japanese MSM with combination interventions. Lancet Reg. Health – West. Pac. 23. https://doi.org/10.1016/j.lanwpc.2022.100467

Wright, S.J., 2015. Coordinate descent algorithms. Math. Program. 151, 3–34. https://doi.org/10.1007/s10107-015-0892-3

Wu, Y., Huang, M., Wang, X., Li, Y., Jiang, L., Yuan, Y., 2020. The prevention and control of tuberculosis: an analysis based on a tuberculosis dynamic model derived from the cases of Americans. BMC Public Health 20, 1173. https://doi.org/10.1186/s12889-020-09260-w

Xue, L., Jing, S., Wang, H., 2022. Evaluating Strategies For Tuberculosis to Achieve the Goals of WHO in China: A Seasonal Age-Structured Model Study. Bull. Math. Biol. 84, 61. https://doi.org/10.1007/s11538-022-01019-1

Yang, C., Kang, J., Lu, L., Guo, X., Shen, X., Cohen, T., Menzies, N.A., 2021. The positive externalities of migrant-based TB control strategy in a Chinese urban population with internal migration: a transmission-dynamic modeling study. BMC Med. 19, 95. https://doi.org/10.1186/s12916-021-01968-9

Zang, X., Krebs, E., Min, J.E., Pandya, A., Marshall, B.D.L., Schackman, B.R., Behrends, C.N., Feaster, D.J., Nosyk, B., Localized HIV Modeling Study Group, 2020. Development and Calibration of a Dynamic HIV Transmission Model for 6 US Cities. Med. Decis. Mak. Int. J. Soc. Med. Decis. Mak. 40, 3–16. https://doi.org/10.1177/0272989X19889356

Zwick, E.D., Pepperell, C.S., Alagoz, O., 2021. Representing Tuberculosis Transmission with Complex Contagion: An Agent-Based Simulation Modeling Approach. Med. Decis. Mak. Int. J. Soc. Med. Decis. Mak. 41, 641–652. https://doi.org/10.1177/0272989X211007842
