## Supplementary figures for "Calibration of transmission-dynamic infectious disease models: a scoping review and reporting framework"

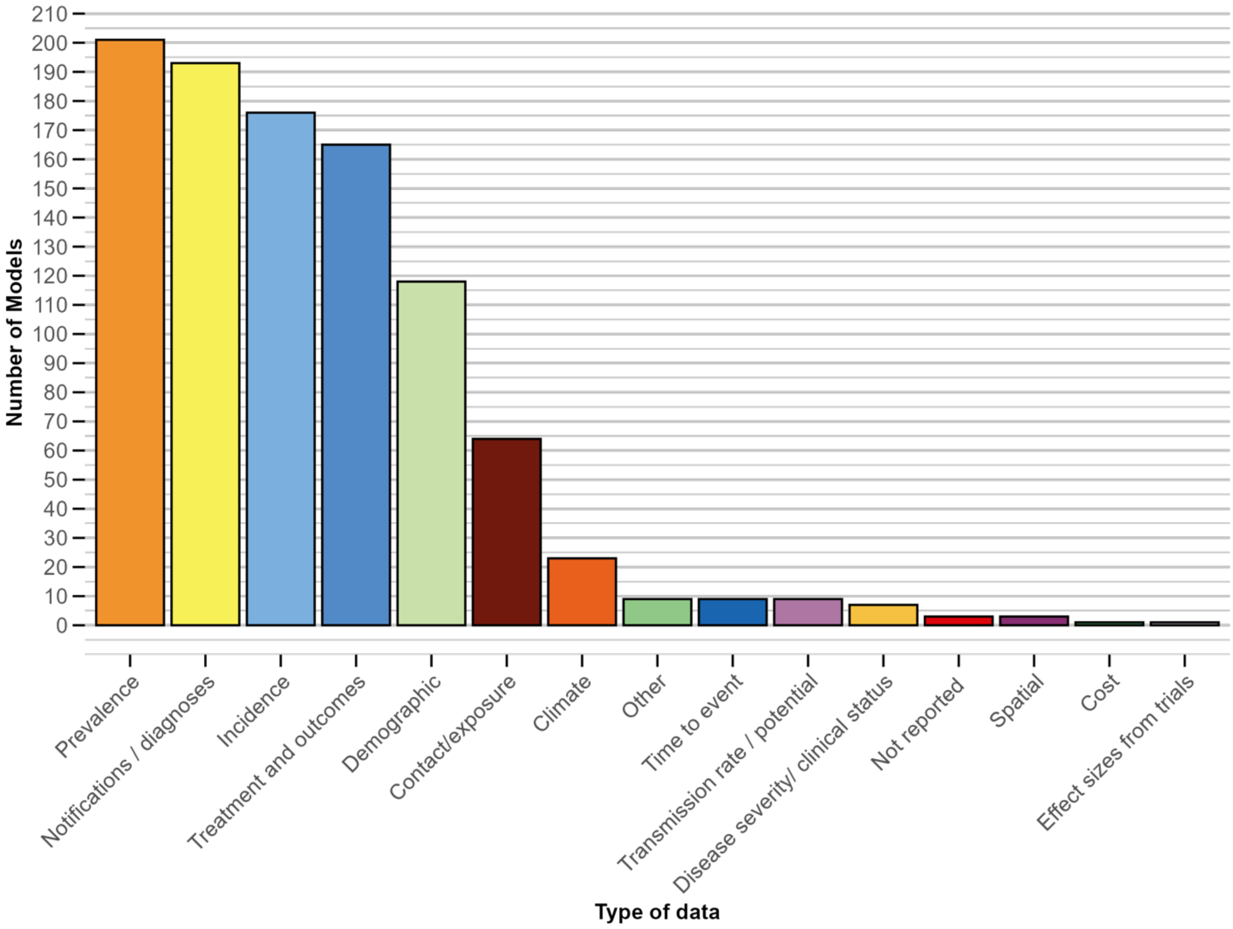


**Figure S1: Frequency of data types used for defining calibration targets.**

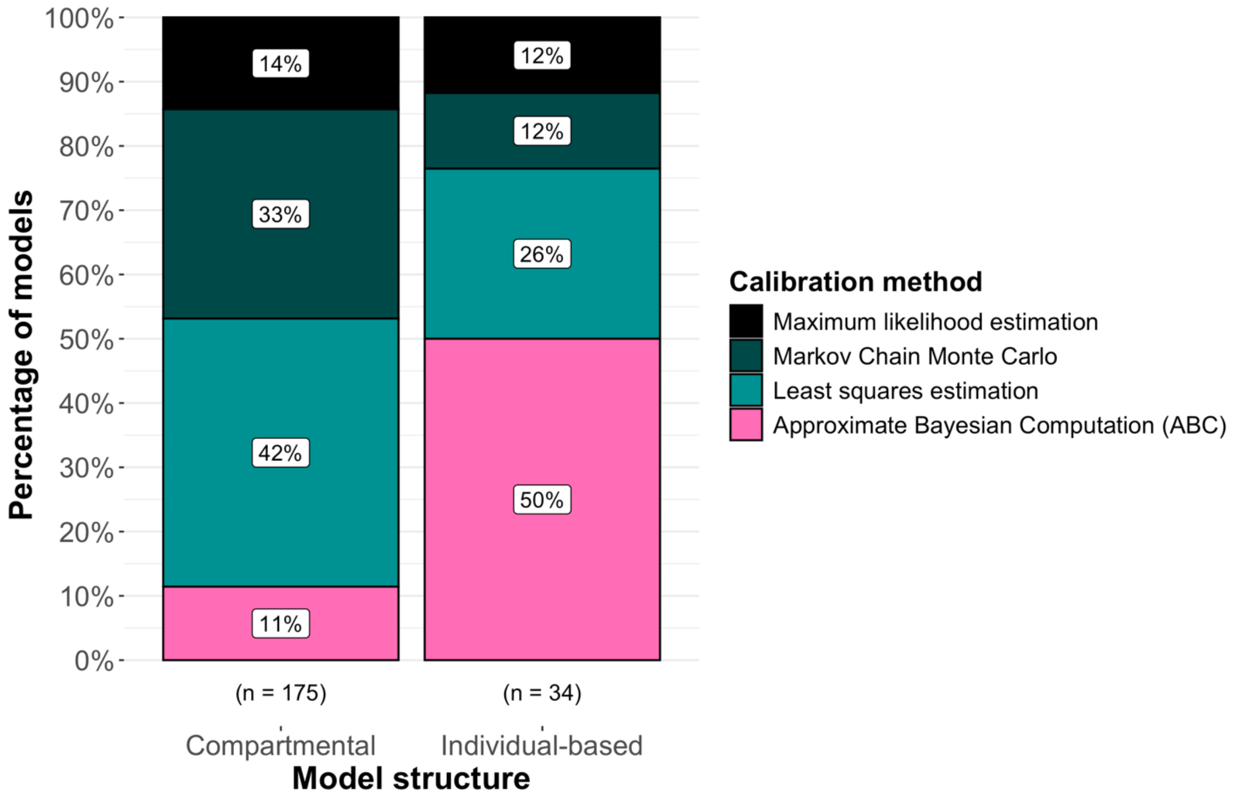


**Figure S2: Most frequent calibration methods (used in at least 30 models) and associated model structure.**

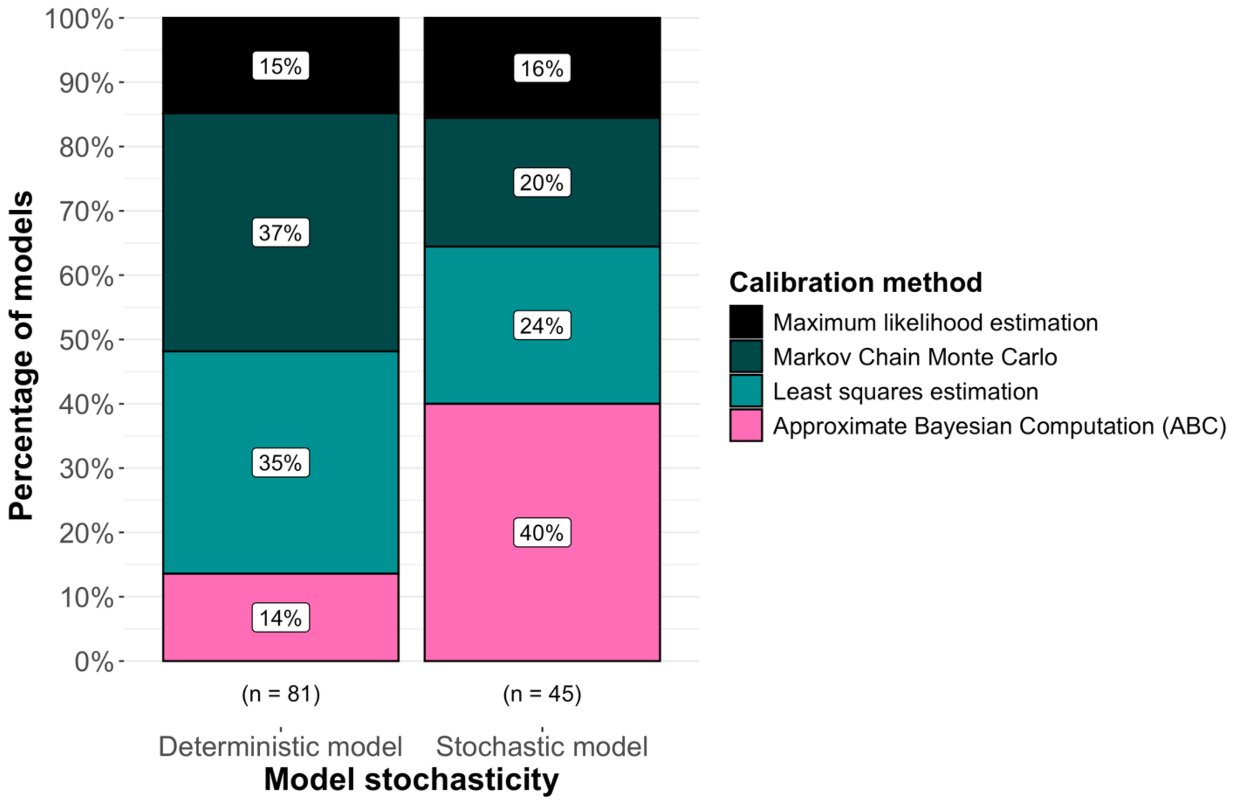


**Figure S3: Most frequent calibration methods (used in at least 30 models) and associated model stochasticity.**

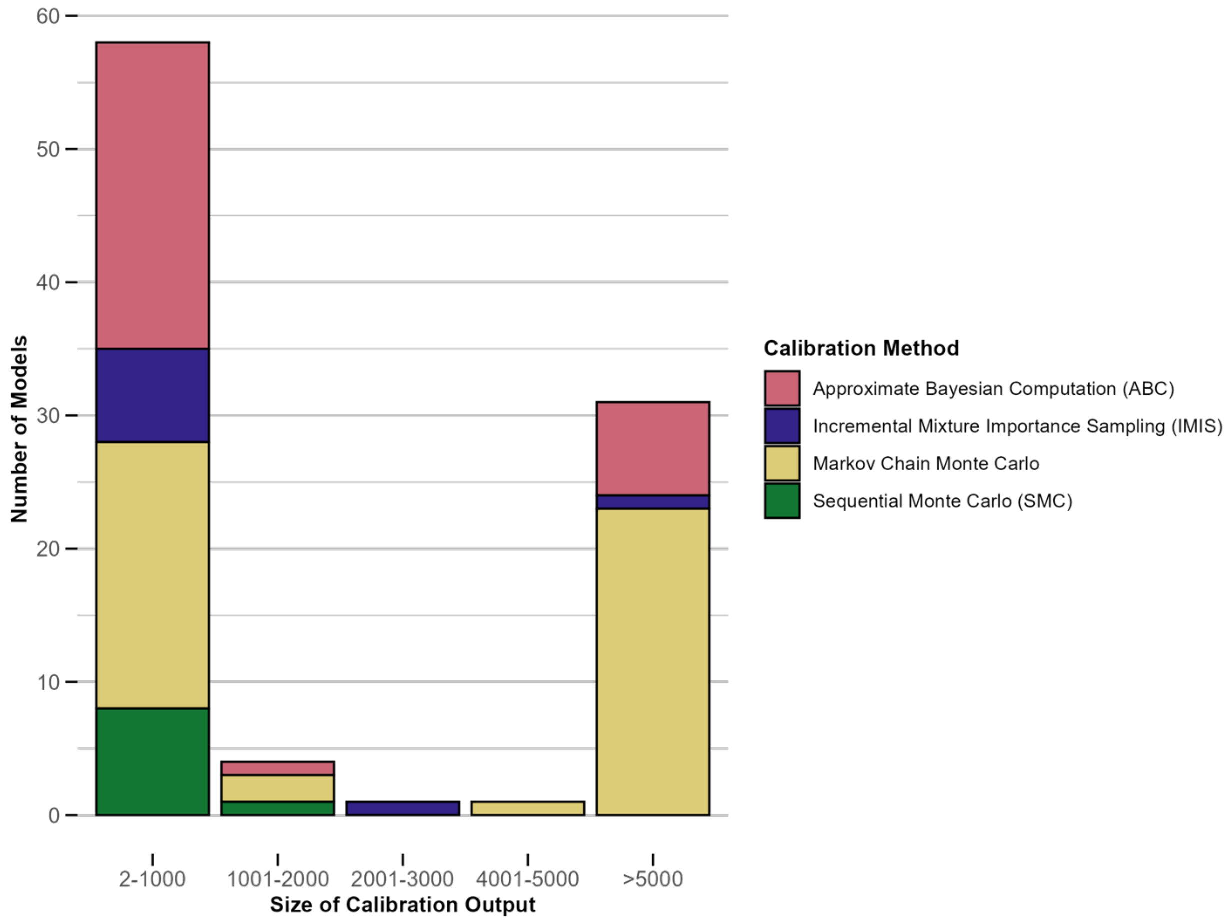


**Figure S4: Distribution of the size of calibration output for outputs greater than size 1; i.e., sample estimates. Results are shown for calibration methods reported in at least 10 models.**

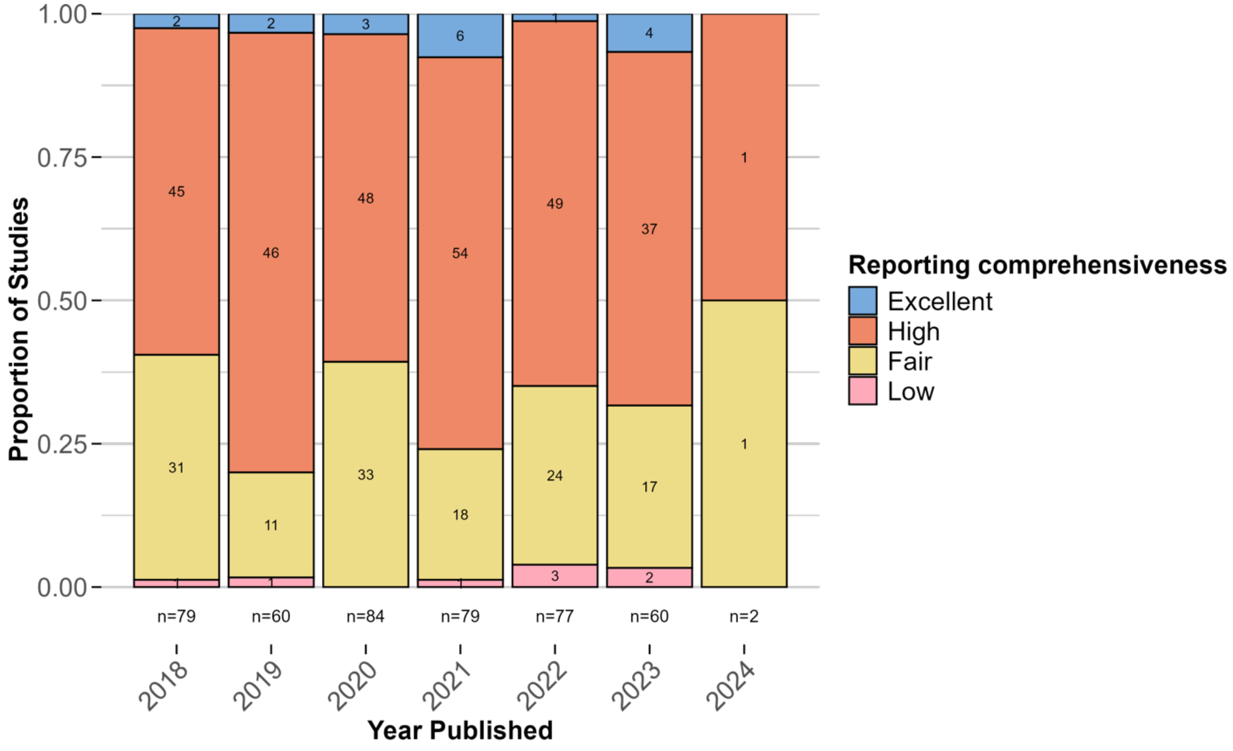


**Figure S5: Distribution of reporting comprehensiveness by year of model publication.**
