## Supplementary material for "Calibration of transmission-dynamic infectious disease models: a scoping review and reporting framework": PIPO reporting framework

### PIPO (Purpose-Inputs-Process-Outputs) framework for calibration reporting

Characteristics of calibration methods are classified into four broad components:

1) **Purpose** (*what is the goal of calibration?*),
2) **Inputs** (*what are the inputs into the calibration algorithm?*),
3) **Process** (*how is calibration conducted, given inputs?*), and
4) **Outputs** (*what are the characteristics of the calibration outputs?*).

**Table 1: PIPO framework in detail.** Classes describe how calibration methods may differ by pillar. Classes are not necessarily mutually exclusive. Refer to accompanying article for details.

| Component | Item # | Possible classes | If reported, page # | Reason for non-reporting, if applicable |
|---|---|---|---|---|
| **A.** ***Purpose: what is the goal of calibration?*** | | | | |
| **A.1. Scientific problem being solved (what is the purpose of calibration?)** | 1 | A.1.1. Understand disease mechanisms (e.g., through inference on a parameter) | | |
| | | A.1.2. Evaluate interventions | | |
| | | A.1.3. Predict disease trends | | |
| | | A.1.4. Assess impact of model assumptions | | |
| **B.** ***Inputs: what are the inputs of the calibration algorithm?*** | | | | |
| **B.1. Parameter inputs** | | | | |
| **B.1.1. Beliefs or evidence** | 2 | B.1.1.1. Prior knowledge is incorporated for at least one parameter to be calibrated; e.g., a prior distribution over parameters which may be sourced from the literature or | | |

| | | |
|---|---|---|
| | | expert judgement (Swallow *et al.*, 2022). In this case, provide references. |
| | | B.1.1.2. No prior knowledge is incorporated for parameters to be calibrated. |
| **B.1.2. Parameters to calibrate.** *Explicitly report which parameters are to be calibrated.* | 3 | B.1.2.1. All parameters |
| | | B.1.2.1. A subset of parameters. |
| **B.1.3. Justification for choice of parameters to calibrate** | 4 | B.1.3.1. Parameters are relevant to question of interest. |
| | | B.1.3.2. Parameters are uncertain. |
| **B.2. Calibration targets** | | |
| **B.2.1. Type of data/estimates used for defining calibration targets.** | 5 | Examples of data or estimate types include incidence, contact/exposure, demography, notifications/diagnoses, prevalence, spatial variables or treatment outcomes.<br><br>*Provide brief descriptions and references for the data or estimate.* |
| **B.2.2. Resolution of data/estimates used for defining calibration targets** | 6 | B.2.2.1. Empirical data or statistical summaries of empirical data (e.g., WHO case notification data) |
| | | B.2.2.2. Modeled estimates (e.g., WHO TB incidence estimates) |
| **B.2.3. Number of calibration targets** | 7 | Example: calibration was performed using five calibration targets |
| **C. *Process: how is calibration conducted, given inputs?*** | | |

| | | |
|---|---|---|
| **C.1. Number of steps** | 8 | C.1.1. Calibration was done as a single step (i.e., all parameters were calibrated at once). |
| | | C.1.2. Calibration was done sequentially: different parameters (or subsets of parameters) were calibrated over multiple rounds |
| **C.2. Name and description of calibration algorithm.** *State and describe, at least briefly, the calibration algorithm employed.* | 9 | Common algorithms include Markov chain Monte Carlo-based methods, Approximate Bayesian Computation and maximum likelihood estimation. |
| **C.3. Calibration implementation.**<br><br>*For more guidance on implementation reporting, refer to Section 4 of the Infectious Disease Modeling Reproducibility Checklist (Pokutnaya et al., 2023).* | 10 | Report an accessible repository for code and relevant dependencies (e.g., data, functions, packages, seed, starting values). Ensure that calibration procedure is: 1) implemented in a clearly stated, accessible programming language or software, 2) implementation is well-documented and has meaningful comments and 3) versions of programming language, package or data repository. |
| **C.4. Goodness-of-fit (GOF) measure employed within calibration algorithm.**<br><br>How is *the level of agreement between modeled outcomes and* | 11 | C.4.1. Ad-hoc distance function (as in Approximate Bayesian Computation or least squares) |
| | | C.4.2. Data likelihood |

| | | |
|---|---|---|
| *calibration targets measured?* | | |
| **D.** ***Output: what are the characteristics of the calibration outputs?*** | | |
| **D.1. Nature of calibration output** | 12 | D.1.1. Point estimate (single parameter value/ single parameter set) |
| | | D.1.2. Sample estimate (multiple parameter values/ multiple parameter sets) |
| | | D.1.3. Distribution estimate (a distribution function which generates parameter values, as obtained with variational inference or Laplace approximation). |
| **D.2. Reporting of calibration outputs** | 13 | D.2.1. Numerical: e.g. as value(s) in a table |
| | | D.2.2 Graphical: e.g., as a plot of calibrated model trend versus calibration targets |
| **D.3. Reporting of uncertainty in calibration output** | 14 | D.3.1. Numerical: As uncertainty intervals around estimates or model outputs. e.g., 95% CI: 1.5 - 4.5 |
| | | D.3.2. Graphical: As a plot, e.g., a line with shaded areas indicating uncertainty intervals |
| **D.4. Size of calibration output**<br><br>*Report the number of parameter sets/ parameter values in calibration output.* | 15 | For example, 1000 parameter sets were produced from the calibration. |

From: Dankwa EA et al. “Calibration conduct and reporting in infectious disease dynamic transmission models: a scoping review and reporting framework”.

**References**

Pokutnaya, D. *et al.* (2023) ‘An implementation framework to improve the transparency and reproducibility of computational models of infectious diseases’, *PLOS Computational Biology*, 19(3), p. e1010856. Available at: https://doi.org/10.1371/journal.pcbi.1010856.

Swallow, B. *et al.* (2022) ‘Challenges in estimation, uncertainty quantification and elicitation for pandemic modelling’, *Epidemics*, 38, p. 100547. Available at: https://doi.org/10.1016/j.epidem.2022.100547.
